# Changes in antimicrobial susceptibility after six months of doxycycline post-exposure prophylaxis: results from a single center, open label, uncontrolled clinical trial (the SafeDoxyPEP trial)

**DOI:** 10.64898/2026.11.18.26368296

**Authors:** Thibaut Vanbaelen, Irith De Baetselier, Sheeba Santhini Manoharan-Basil, Said Abdellati, Vicky Cuylaerts, Ilse Tack, Yven Van Herrewege, Lida van Petersen, Soumia El Maadouri, Edith Kitoko, Kaat Vanderheyden, Lindsay Poppe, Kristien Wouters, Tessa De Block, Dorien Van Den Bossche, Achilleas Tsoumanis, Chris Kenyon

**Affiliations:** Department of Clinical Sciences, Institute of Tropical Medicine, Antwerp 2000, Belgium; Division of Infectious Diseases and HIV Medicine, University of Cape Town, Cape Town 7700, South Africa

## Abstract

**Synopsis:** 

**Background:** Doxycycline post-exposure prophylaxis (doxyPEP) reduces the incidence of bacterial sexually transmitted infections, but its impact on antimicrobial susceptibility among bystander bacteria remains unclear.

**Objectives:** To assess changes in susceptibility to doxycycline among rectal *Escherichia coli* after 6 months of doxyPEP. Secondary objectives assessed susceptibility for other classes of antimicrobials and in other species.

**Patients and methods:** Single-centre, open-label, single-arm trial at an HIV pre-exposure prophylaxis clinic in Belgium. Participants used doxycycline 200mg within 72h after condomless sex for 180 days. Anorectal swabs and oral rinse samples were collected at baseline and day 180. Susceptibility was assessed by disc diffusion on six colonies per species. Differences in inhibition-zone diameters were analysed using censored Bayesian mixed-effects Gaussian regression.

**Results:** Seventy-nine participants were enrolled, 73 completed day 180. Among rectal isolates, cefixime inhibition-zone diameters decreased in *E. coli* and *Klebsiella* spp., other estimates were imprecise. Among oral isolates, inhibition-zone diameters decreased for doxycycline and azithromycin in streptococci, staphylococci and commensal *Neisseria* spp.; ceftriaxone and cefixime in commensal *Neisseria* spp.; and ciprofloxacin and penicillin in staphylococci and commensal *Neisseria* spp. Ceftriaxone inhibition-zone diameters increased in oral streptococci. Sensitivity analyses were broadly consistent. Twenty-five participants experienced drug-related adverse events, none serious.

**Conclusions:** Six months of doxyPEP was associated with reduced susceptibility to doxycycline and several other antimicrobials, predominantly among oral bacteria. Rectal findings were less precise apart from reduced cefixime susceptibility in *E. coli* and *Klebsiella* spp. These findings support further studies of DoxyPEP and bystander selection across multiple bacterial taxa and antimicrobial classes.

## Introduction

Doxycycline post exposure prophylaxis (doxyPEP) reduces chlamydia and syphilis incidence among men who have sex with men, and several trials have also reported protection against gonorrhoea.^1–4^ Repeated antimicrobial exposure may, however, affect organisms other than the pathogens that prophylaxis is intended to prevent.^2,5^ Bystander selection is relevant because commensal and potentially pathogenic bacteria can act as reservoirs of reduced susceptibility, and selection involving one antimicrobial class may result in changes in susceptibility to other classes.^6–9^

Early doxycycline PEP studies were designed primarily to estimate efficacy against sexually transmitted infections (STI). Their microbiological analyses often relied on small isolate numbers or binary susceptibility categories. A number of studies have found that dichotomising a continuous measure of antimicrobial susceptibility into resistant and susceptible can obscure distributional shifts in antimicrobial susceptibility.^3,10–12^ SafeDoxyPEP therefore examined quantitative susceptibility measures in bacteria from the rectum and oropharynx following six months of doxycycline PEP. The study also expanded the range of bacteria-antimicrobial combinations assessed. A number of studies have found that tetracycline resistance is strongly associated with resistance to other classes of antimicrobials in a range of bacterial species.^13–15^ Adding to this concern, one doxyPEP randomized controlled trial (RCT) found that doxyPEP was associated with selection of gonococci with reduced susceptibility to cefixime.^16^ A recent observational study also found an association between doxyPEP use and selection for tetracycline resistance in *Staphylococcus aureus* and Group A Streptococcus (GAS).^17^

The primary objective was to assess changes in doxycycline susceptibility among rectal *Escherichia coli* between baseline and day 180 of doxyPEP use. Secondary objectives included assessing changes in susceptibility to azithromycin, doxycycline, cefixime, ciprofloxacin, ceftriaxone and penicillin among rectal *E. coli* and *Klebsiella* spp., as well as oral streptococci, staphylococci and commensal *Neisseria* spp. Organism–antimicrobial combinations with intrinsic resistance were excluded from the analyses.

## Patients and methods

### Study design and participants

SafeDoxyPEP was a prospective, single-centre, single-arm, open-label trial at the HIV pre-exposure prophylaxis (PrEP) clinic of the Institute of Tropical Medicine, Antwerp, Belgium. Participants were recruited during routine PrEP visits. Eligible participants were adults, assigned male at birth, eligible for HIV PrEP, able to consent and willing to follow study procedures. Principal exclusions were antibiotic use during the preceding three months, contraindication or allergy to doxycycline or product excipients, and use of a vitamin K antagonist.

Participants were asked to take two tablets of doxycycline monohydrate 100 mg ideally within 24 h and no later than 72 h after condomless anogenital, vaginal or oral sex, with no more than one dose per 24 h. Follow-up visits were scheduled at days 90 and 180. Adherence was assessed by self-report and returned-tablet counts.

### Laboratory procedures and outcomes

At each study visit, participants self-collected an oral rinse and two anorectal E-swabs. For the oral rinse, participants rinsed their mouth with 15 mL of bottled water for 30 s and expelled the fluid into a container. For anorectal sampling, swabs were inserted approximately 5 cm into the anus and rotated three times; the swab intended for culture was collected first.

All samples were stored at -80°C until batch processing at the end of the study. For oral-rinse processing, three aliquots were prepared by adding 750 µL of oral rinse to 750 µL of skim milk cryopreservation medium containing 30% glycerol. For the culture anorectal swab, two aliquots were prepared from the primary E-swab tube by mixing 750 µL of E-swab transport medium with 750 µL of skim milk containing 30% glycerol; after vortexing, 750 µL of the mixture was transferred to a second cryotube, and both aliquots were stored at -80°C.

Rectal samples were cultured for *E. coli* and *Klebsiella spp*., and oral-rinse samples for streptococci, staphylococci and commensal *Neisseria* spp.

Selective agar media were used to isolate the target organisms as follows:

#### E. coli

Chromocult Coliform Agar (Sigma-Aldrich, Merck) was used for selective differentiation of coliforms. *E. coli* are the only bacteria to appear as blue colonies, whereas other coliforms appear as pink colonies. 100 µL of undiluted stored E-swab sample was plated per plate.

#### Klebsiella spp

Klebsiella ChromoSelect Selective Agar (KCSA) (Sigma-Aldrich, Merck) with the addition of Klebsiella Selective Supplement (Sigma-Aldrich, Merck) was used for culture. *Klebsiella spp*. are the only bacteria to grow as pink colonies on this media. 100 µL of undiluted stored E-swab sample was plated onto each agar plate.

#### Neisseria spp

Luria-Bertani-medium supplemented with Vancomycin and Trimethoprim (LBVT.SNR) was used for culture. 100 µL of undiluted stored oral rinse sample was plated per plate.

#### Streptococci spp

Columbia Nalidixic agar (CNA, Thermo Fisher Scientific) was used for culture. 100 µL of a 1:100 dilution of the stored oral rinse was plated per plate.

#### Staphylococci spp

Mannitol Salt Agar (MSA) (Thermo Fisher Scientific) was used for culture. 100 µL of undiluted stored oral rinse was plated per plate.

All plates were incubated for 24-48 hours in a CO_2_ incubator at 36.5°C and 5.5% CO_2_ to promote bacterial growth.

Six colonies with expected morphology were randomly selected from each plate of each species. Antibiotic susceptibility (azithromycin, doxycycline, cefixime, ceftriaxone, ciprofloxacin, benzylpenicillin) was assessed via the Kirby-Bauer disc diffusion method, according to EUCAST guidelines.^4^ The disc diameter was 6mm.

The primary outcome was the change from baseline to day 180 in doxycycline susceptibility of rectal *E. coli*. Secondary outcomes were corresponding changes for azithromycin, cefixime, ceftriaxone and ciprofloxacin in *E. coli* and in *Klebsiella spp*.; and the same four antimicrobials plus doxycycline and penicillin in oral streptococci, staphylococci and commensal *Neisseria* spp. Organism-antimicrobial combinations with intrinsic or expected resistance were excluded from the analysis. Safety outcomes comprised solicited and unsolicited adverse events, seriousness and investigator-assessed relatedness.

### Comparison of disc diffusion diameters with MICs

Eighty-six colonies picked from the chromogenic *E. coli* medium at baseline, from 16 participants, had MICs determined for doxycycline, ceftriaxone and ciprofloxacin in addition to disc diffusion. Of these, 84 were identified as *E. coli*, one as *Klebsiella oxytoca* and one as *Pantoea septica*. No month-6 isolates and no isolates from the other four selective media were tested by both methods, so the calibration is confined to baseline *E. coli*.

For each antimicrobial, log2(MIC) was regressed on zone diameter by ordinary least squares, and the reciprocal of the slope taken as the zone difference corresponding to a twofold change in MIC. Observations at an assay limit were retained in the primary fit and identified separately: 12 doxycycline and 3 ciprofloxacin zones sat at the 6 mm disc diameter, and 2 doxycycline, 11 ceftriaxone and 3 ciprofloxacin MICs sat at the highest concentration tested. The doxycycline regression was repeated excluding every observation at either limit.

### Assessing for a secular trend in antimicrobial susceptibility in baseline visits

A reduction in zone diameters at month 6 could in principle reflect something that changed in the laboratory or in the source population over the study period rather than anything doxyPEP did: a new lot of agar or of antimicrobial discs, a change of reader, seasonal variation in carriage, or secular change in who was enrolling.

Because enrolment to this trial was staggered over eight months, we could test for this in two independent ways. Test 1: secular trend among pre-treatment specimens. Test 2: comparison within a common laboratory window. For details see SBox 1.

### Sample size

The sample size was calculated by simulating a Wilcoxon signed rank test on paired data to find a power of 81%. The assumed effect size was based on the paper by Harrison et al who reported a difference of 0.6 among 62 results of MIC to tetracycline for gonorrhea cases ^18^. The expected power is at least 80% at a 5% significance level. The calculated sample size is 70 participants, which was corrected for an expected 10% attrition rate, resulting in a final sample size of 78 participants.

### Statistical analysis

The intention-to-treat approach was the primary one, according to GCP-E9 guidelines for superiority studies. Participants were included in an anatomical sampling group if they had a susceptibility measurement at baseline and day 180. The per-protocol population excluded major eligibility or treatment deviations, no doxycycline PEP use, non-study antibiotic use, visits outside the permitted window or failure to provide samples. Participants taking at least one dose formed the safety population.

Repeated colony measurements were analysed using censored Bayesian mixed-effects Gaussian regression. The smallest diameter that could be measured on the antimicrobial discs was 6 mm (diameter of the disc), thus results with value 6mm were treated as left-censored. Visit was a categorical fixed effect. Models included participant-specific random intercepts and random slopes for visit. The day-180 coefficient estimates the average change in inhibition-zone diameter, with 95% credible intervals (CrIs). Separate models were fitted for each organism-antibiotic combination. Models were repeated in the per-protocol population and adjusted separately for cumulative doxycycline tablet use and non-study antibiotic exposure. The analyses were repeated in a subset of participants reporting high consumption of doxycycline. High consumption was defined as ≥ 27 tablets of doxycycline during the study. A sensitivity analysis was performed for the primary and secondary outcomes replacing the smallest diameter of inhibition measured (6mm) with 1mm. Analyses were performed using R version 4.5.2.

The study is reported in line with the Transparent Reporting of Evaluations with Nonrandomised Designs (TREND) statement.^19^

### Ethics

The trial was approved by the relevant Belgian competent authority and ethics committee (EU-CT 2023-507137-24-00). All participants provided written informed consent. Conduct followed the Declaration of Helsinki, Good Clinical Practice and Good Clinical Laboratory Practice.

## Results

### Participants and exposure

Seventy-nine participants were enrolled between March 2024 and June 2025 (Figure 1). Three were excluded from all analyses, leaving 76 participants; 73 completed day 180. Fifty-five participants were included in rectal analyses and 73 in oral analyses. The per-protocol population comprised 31 participants, and 76 were included in safety analyses.

**Figure 1.**
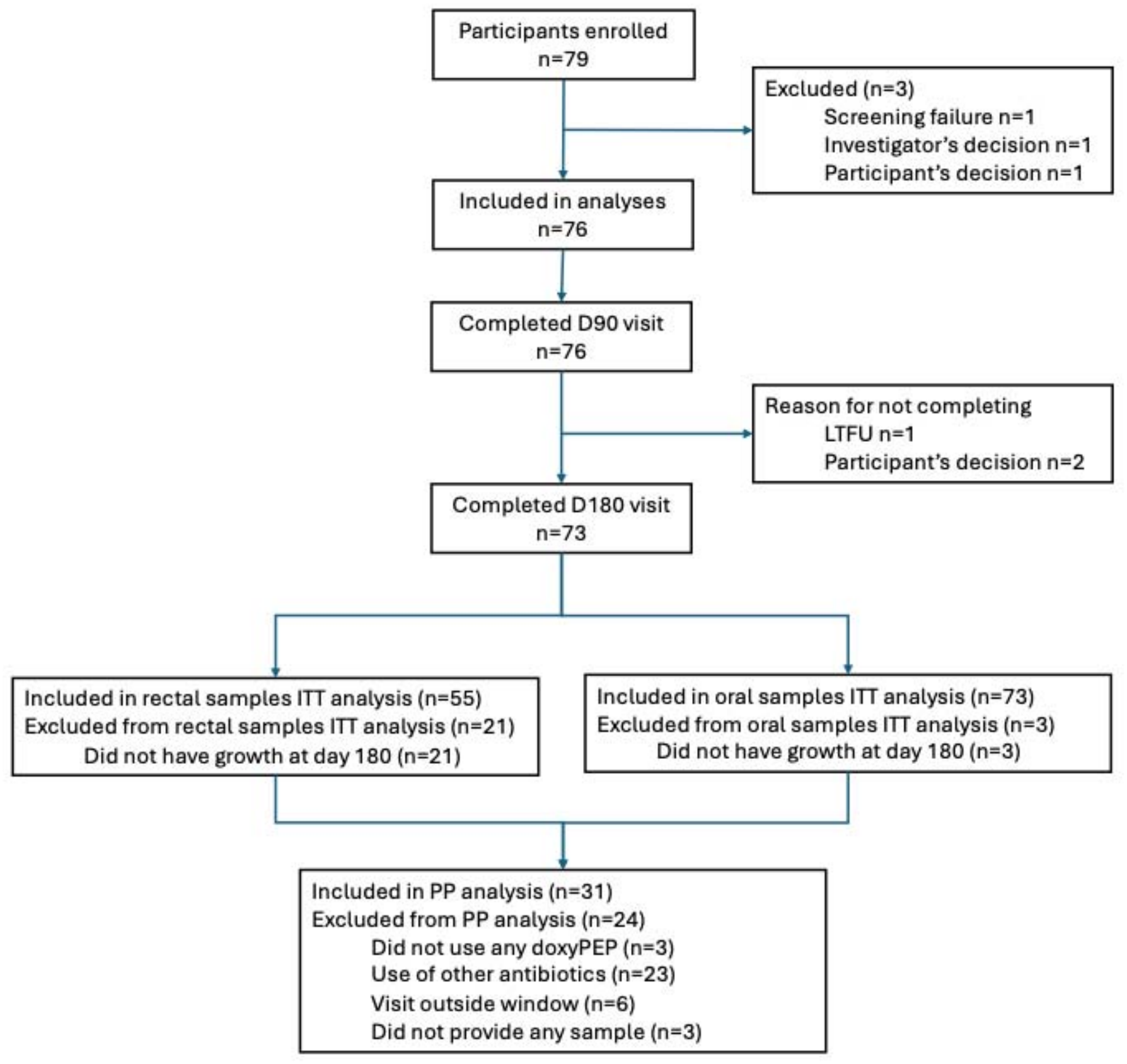
Study flowchart.

All participants were male at birth (Table 1). Median age was 42 years (IQR 36–48.25), and 26 (34.2%) reported a bacterial STI in the prior 6 months. Participants reported a median of 15 (IQR 8–25) partners in the prior 3 months (SFigure 4). Median reported doxycycline use was 19 tablets (IQR 10–29.25) from baseline to day 90 and 23 tablets (IQR 15–37) from day 90 to day 180 (SFigure 1). Condom use during anal sex was generally low, and a minority reported less condom use after starting doxycycline PEP (SFigures 2 and 3).

**Table 1.** Baseline descriptive demographic and behavioural characteristics.

| **Characteristic** | **n (%) or median (IQR)** |
|---|---|
| **Sex: male** | 76 (100) |
| **Age, years** | 42 (36–48.25) |
| **Sex partners in previous 3 months** | 15 (8–25) |
| **Mouthwash use in previous month** | 39 (51.3) |
| **Any bacterial STI in previous 6 months\*** | 26 (34.2) |
| **Chlamydia** | 8 (10.5) |
| **Gonorrhoea** | 11 (14.5) |
| **Syphilis** | 11 (14.5) |
| **Unknown STI type** | 1 (1.3) |
| **Any antibiotic in last 24 months** | 56 (73.7) |

\* There were participants with multiple bacterial STIs, thus the individual counts are greater than the overall sum.

**Figure 2.**
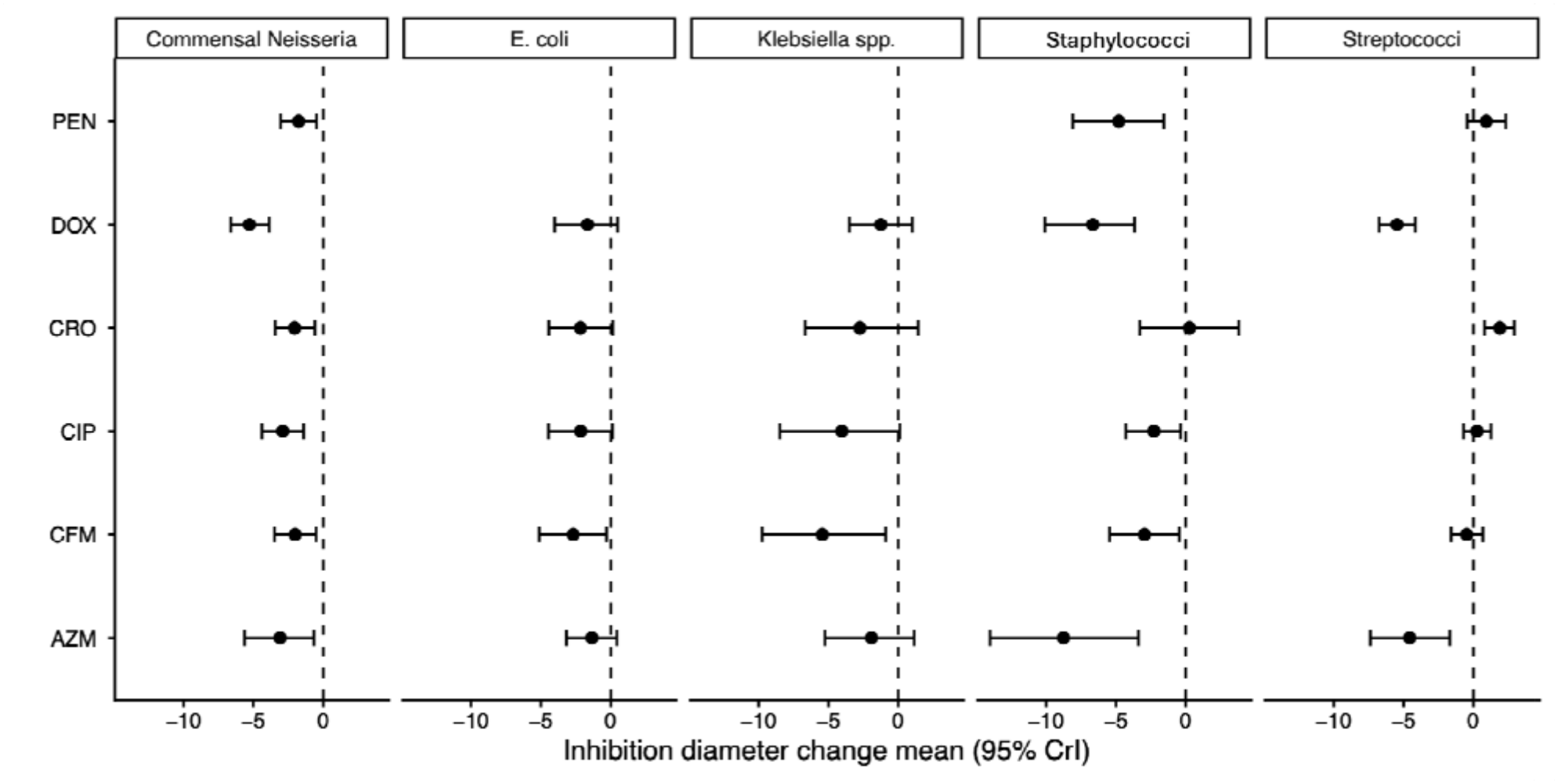
Estimated change from baseline to day 180 in inhibition-zone diameter, intention-to-treat analysis. (PEN – penicillin, DOX – doxycycline, CRO – ceftriaxone, CIP – ciprofloxacin, CFM – cefixime, AZM – azithromycin)

### Rectal susceptibility outcomes

In the intention-to-treat analysis, changes in rectal inhibition-zone diameters between baseline and day 180 were generally modest, with most 95% CrIs including zero (Figure 2). For the primary endpoint, the inhibition-zone diameter for doxycycline in rectal *E. coli* decreased by an estimated -1.67 mm (95% CrI -4.02 to 0.49).

There were statistically significant declines in inhibition-zone diameters of -2.69 mm (95% CrI - 5.12 to -0.29) for cefixime in rectal *E. coli* and -5.44 mm (95% CrI -9.75 to -0.90) for cefixime in rectal *Klebsiella spp*.

Per-protocol analyses were directionally similar, with a larger and statistically significant decline in *E. coli* doxycycline susceptibility (-3.23 mm, 95% CrI -6.31 to -0.04; STable 5).

Sensitivity analyses that handled values at the lower boundary differently and were stratified by doxycycline consumption yielded similar inferences for the principal outcomes (STables 1,3,11).

Between-participant heterogeneity was substantial throughout, with random-slope standard deviations ranging from approximately 6 to 14 mm in the intention-to-treat rectal models (Figure 2; Stable 13).

### Oral susceptibility outcomes

Among oral isolates, the inhibition zone for doxycycline declined consistently across all three organism groups: -5.47 mm (95% CrI -6.75 to -4.15) in streptococci, -6.66 mm (95% CrI -10.09 to -3.67) in staphylococci and -5.28 mm (95% CrI -6.62 to -3.86) in commensal *Neisseria* spp. (Figure 2).

For cefixime, declines were significant in staphylococci (-2.96 mm [95% CrI -5.46 to -0.47]) and commensal *Neisseria* spp. (-2.01 mm [95% CrI -3.49 to -0.51]).

For ceftriaxone, inhibition-zone diameters increased in oral streptococci by 1.89 mm (95% CrI 0.81 to 2.93), were unchanged in staphylococci, and declined in commensal *Neisseria* spp. by - 2.05 mm (95% CrI -3.43 to -0.60).

Ciprofloxacin susceptibility declined significantly in staphylococci (-2.29 mm [95% CrI -4.30 to - 0.38]) and commensal *Neisseria* spp. (-2.90 mm [95% CrI -4.39 to -1.40]).

Azithromycin susceptibility declined significantly in all three oral organism groups: -4.56 mm (95% CrI -7.37 to -1.68) in streptococci, -8.76 mm (95% CrI -14.02 to -3.39) in staphylococci and -3.08 mm (95% CrI -5.63 to -0.66) in commensal *Neisseria* spp.

Penicillin susceptibility declined in staphylococci (-4.80 mm [95% CrI -8.10 to -1.58]).

Per-protocol and sensitivity analyses were generally consistent, with larger-magnitude declines for several outcomes in oral staphylococci (STables 2,4,6,12).

**Table 2.** Summary of safety endpoints, n/N (%; 95% CI)

| **AE** | **Overall** | **Day 90** | **Day 180** |
|---|---|---|---|
| **Death** | 0/76 (0; 0–4.8) | 0/76 (0; 0–4.8) | 0/73 (0; 0–5.0) |
| **Serious AE** | 2/76 (2.6; 0.7–9.1) | 2/76 (2.6; 0.7–9.1) | 1/73 (1.4; 0.2–7.4) |
| **Drug-related serious AE** | 0/76 (0; 0–4.8) | 0/76 (0; 0–4.8) | 0/73 (0; 0–5.0) |
| **Drug-related AE** | 25/76 (32.9; 23.4–44.1) | 15/76 (19.7; 12.3–30.0) | 14/73 (19.2; 11.8–29.7) |
| **Any AE** | 42/76 (55.3; 44.1–65.9) | 26/76 (34.2; 24.5–45.4) | 26/73 (35.6; 25.6–47.1) |

AE, adverse event.

### Safety

Forty-two of 76 treated participants (55.3%) experienced at least one adverse event and 25 (32.9%) experienced an event assessed as drug related (Table 2). Diarrhoea occurred in 10 (13.2%), dyspepsia in eight (10.5%), nausea in six (7.9%), and photosensitivity reaction and rash in two participants each (2.6%). Two participants experienced serious adverse events, neither related to doxycycline. There were no deaths and no drug-related serious adverse events.

### Secular trend in antimicrobial susceptibility

We found no evidence of a reduction in antimicrobial susceptibility over time (SBox 1).

### Disc diffusion versus MICs

Zone diameter and log2(MIC) were closely and linearly related (SBox 2). For doxycycline the slope was -0.320 log2 units per millimetre (95% CI -0.350 to -0.289; r = -0.92, R-squared = 0.84), so 3.13 mm of zone diameter corresponds to a twofold change in MIC. Excluding all observations at an assay limit gave a nearly identical 3.05 mm per doubling (n = 73, R-squared = 0.82), so the relationship is not an artefact of the censored extremes. Ceftriaxone was steeper, at 1.91 mm per doubling (R-squared = 0.89), and ciprofloxacin shallower and considerably noisier, at 3.47 mm per doubling (R-squared = 0.57).

## Discussion

In this six-month single-arm study of doxycycline PEP, the most consistent susceptibility changes were observed in oral bacteria rather than rectal Enterobacterales. Oral streptococci, staphylococci and commensal *Neisseria* spp. all showed reduced doxycycline susceptibility by day 180, and several non-doxycycline agents also showed declines, particularly azithromycin and, for selected organisms, cefixime, ceftriaxone, ciprofloxacin and penicillin. In contrast, rectal analyses were generally imprecise, with the clearest signal being reduced cefixime susceptibility in both *E. coli* and *Klebsiella* spp.

These findings support the concern that repeated doxycycline exposure may co-select for altered susceptibility beyond tetracyclines. The oral compartment may be especially vulnerable because it contains diverse, exchange-prone commensal communities. The pattern of declines in azithromycin, cefixime, ceftriaxone and penicillin susceptibility is consistent with the possibility of broader ecological selection or strain replacement rather than an isolated effect on doxycycline susceptibility alone.

The cephalosporin findings merit cautious interpretation. In rectal isolates, mean susceptibility to ceftriaxone and cefixime tended to decline, although the estimates were imprecise. In the oral compartment, reduced susceptibility to both agents was observed in commensal *Neisseria* spp., whereas oral streptococci showed increased ceftriaxone susceptibility and little evidence of change in cefixime susceptibility. A directly comparable pattern in oral streptococci has not, to our knowledge, been reported in other clinical doxycycline PEP trials. Nevertheless, doxycycline and other tetracycline exposures can select resistant organisms within the oropharyngeal microbiota without producing major changes in overall microbial diversity, and may permit ecological replacement of susceptible strains or species by resistant lineages.^20^

The increase in ceftriaxone susceptibility among oral streptococci may therefore reflect altered species or strain composition rather than a genuine within-strain increase in susceptibility. In particular, selection of tetracycline-resistant streptococcal lineages that happen to retain greater ceftriaxone susceptibility could produce the observed results.^21,22^ One observational study found that doxyPEP was associated with an increased prevalence of oral *Streptococcus pyogenes*, which they hypothesized was due in part to the fact 82% of isolates in their study were resistant to tetracyclines.^17^ Another observational study in a PrEP cohort found an association between receipt of doxycycline and decreased oral streptococcal susceptibility to azithromycin.^23^ However, species-level identification and longitudinal genomic comparison of baseline and follow-up isolates would be required to distinguish strain replacement from phenotypic change within persistent colonizing strains.

The oral *Neisseria* findings warrant particular consideration. Commensal *Neisseria* are frequent oropharyngeal colonizers and an established reservoir from which resistance determinants, including mosaic *penA* alleles associated with reduced extended-spectrum cephalosporin susceptibility, can be transferred to *N. gonorrhoeae*.^20,24^ Experimental work has shown that doxycycline exposure can select doxycycline-resistant commensal *Neisseria*, while co-culture experiments demonstrate transfer of mobile tetracycline- and beta-lactam-resistance determinants between gonococci and commensal *Neisseria*.^20,25^ These data do not establish that doxyPEP caused the cephalosporin shifts observed here, but taken in conjunction with the RCT evidence that doxyPEP selected for reduced gonococcal susceptibility to cefixime,^16^ they strengthen the rationale for species-resolved phenotypic and genomic surveillance of the oropharyngeal *Neisseria* reservoir.

Our findings do not demonstrate absence of an effect where intervals crossed the null. The models estimate a population-average change, whereas individual trajectories varied markedly. Opposing changes can yield a small average even when some participants have substantial shifts. Variation may reflect intermittent exposure, strain turnover, acquisition or loss of organisms, colony sampling or measurement variability.

There were several methodological limitations. The study was not randomized and lacked a control group, so secular trends unrelated to doxycycline PEP cannot be excluded. The study was underpowered, especially for rectal analyses, because usable rectal cultures were available for fewer participants than anticipated. The outcome was based on disc-diffusion inhibition zones rather than MICs. Disc diffusion and MICs are related but not interchangeable, and the use of 6 mm discs introduced left-censoring because values could not fall below the physical disc diameter. ^26,27^ If resistant isolates were already clustered at this lower boundary, further phenotypic deterioration would be difficult to detect. A further limitation is that the day-90 samples were not tested, so earlier changes could not be examined.

The study also had important strengths. It used prospective paired sampling, multiple colonies per sample, models that accounted for clustering within participants, and assessment of both rectal and oral bacteria across multiple antimicrobial classes. Follow-up was high, and safety findings were consistent with the known gastrointestinal tolerability profile of doxycycline.^28,29^

In conclusion, six months of doxycycline PEP was associated with measurable reductions in susceptibility for several oral bacteria, affecting doxycycline as well as azithromycin, cefixime, ceftriaxone, ciprofloxacin and penicillin in selected organism-antibiotic combinations. Rectal findings were less consistent, although cefixime susceptibility declined in *E. coli* and *Klebsiella spp*. Larger controlled studies with molecular characterization are needed to determine the mechanisms, persistence and clinical relevance of these shifts. Our findings call for caution regarding the widespread implementation of doxyPEP.

## Supporting information

TREND Checklist

## Data Availability

Anonymized participant-level data may be available through the Institute of Tropical Medicine managed-access procedure, subject to review, a data-sharing agreement and GDPR compliance.

## Acknowledgements

We thank the participants and the clinical, laboratory, data-management and clinical-trial staff at the Institute of Tropical Medicine.

## Transparency declarations

The authors declare no conflicts of interest.

## Author contributions

C.K., T.V., I.D.B. and S.S.M.-B conceived the study. T.V., I.D.B., S.S.M.-B., S.A., V.C., Y.V.H., L.v.P., A.D., K.W., D.V.D.B., S.E.M., E.K., K.V., and C.K. contributed to study implementation, data acquisition or laboratory procedures. A.T. performed the statistical analyses. C.K. drafted the manuscript. All authors critically revised the manuscript, approved the final version and agree to be accountable for the work.

