## Supplementary material for "Changes in antimicrobial susceptibility after six months of doxycycline post-exposure prophylaxis: results from a single center, open label, uncontrolled clinical trial (the SafeDoxyPEP trial)": Online Appendix

**Supplementary appendix**

[Supplementary Figure S4: Number of reported sex partners per study visit (median [thick black line] and IQR [box]).	27](#_Toc243861657)

**Supplementary Methods**

The statistical analysis report included several additional analyses that were not presented in the main manuscript. These comprised sensitivity analyses for alternative handling of lower-bound inhibition values, per-protocol analyses, models adjusted for cumulative doxycycline tablet use and subgroup analyses by level of doxycycline exposure. The main manuscript focused on the intention-to-treat day-180 inhibition-zone analyses for the 6 antimicrobials assessed. The supplementary appendix therefore presents selected additional analyses that are most informative for interpretation of the main findings.

In the primary models, inhibition-zone diameters were analysed using censored Bayesian mixed-effects Gaussian regression with a lower boundary at 6 mm. In sensitivity analyses, values at the lower boundary were alternatively handled by replacing them with 1 mm in Bayesian mixed-effects models. We also fitted exploratory models including cumulative doxycycline tablets over the last 3-month period as an additional fixed covariate.

#### Supplementary Box 1. Secular laboratory drift and seasonality do not explain the reduction in inhibition zone diameters at month 6

###### Rationale

In a single-arm trial, every participant's month-6 specimen is collected six months after their baseline specimen, so the visit contrast is perfectly confounded with elapsed calendar time within any individual. A reduction in zone diameters at month 6 could therefore in principle reflect something that changed in the laboratory or in the source population over the study period rather than anything doxyPEP did: a new lot of agar or of antimicrobial discs, a change of reader, seasonal variation in carriage, or secular change in who was enrolling. This is the most important non-causal explanation for the trial's central finding.

Enrolment to this trial was staggered over eight months, and that makes the objection testable in two independent ways. We present both.

###### Methods

Visit dates were recovered for all 146 study visits. Baseline visits ran from 27 March to 14 November 2024 and month-6 visits from 11 September 2024 to 19 June 2025, with a median interval of 210 days (range 167 to 308).

Test 1: secular trend among pre-treatment specimens. All 73 baseline specimens were collected before any participant had taken doxyPEP, so any association between the calendar date of enrolment and the baseline zone diameter cannot be an effect of the intervention. It can only reflect drift in the measurement system or change in the population being sampled. For each of the 29 plate-by-antimicrobial combinations we regressed the participant mean baseline zone diameter on the ordinal date of enrolment by ordinary least squares, expressing the slope as millimetres per 30-day month, and summarised the direction and significance of the 29 slopes. We separately tested whether the number of antibiotic courses in the 24 months before enrolment, or the number of doxyPEP tablets subsequently taken, varied with enrolment date, to distinguish drift in the laboratory from drift in the cohort.

Test 2: comparison within a common laboratory window. Because enrolment was staggered, the baseline specimens of late-enrolling participants were processed in the same calendar period as the month-6 specimens of early-enrolling participants. A 64-day window from 11 September to 14 November 2024 contains 23 baseline visits and 16 month-6 visits, with no participant contributing both. Comparing the two groups holds calendar time, reagent lots and laboratory conditions fixed by construction; it is a between-participant rather than a paired comparison and is correspondingly less powerful. For each combination we compared participant mean zone diameters between the two groups with the Mann-Whitney test and Welch's t-test, and we compared the resulting 29 differences with the corresponding within-participant paired changes in the full cohort by Spearman correlation. Balance between the two groups was assessed for doxyPEP adherence, prior antibiotic courses and age.

###### Results

Test 1. There was no systematic secular trend in baseline susceptibility. Of the 29 slopes, 12 were negative, which is no more than chance would give, and only 5 reached nominal significance. For doxycycline specifically, the slope was positive in 4 of the 5 taxa and reached significance in none (range -0.36 to +0.46 mm per month). The largest trends ran in the direction opposite to an artefactual explanation, later-enrolling participants having wider rather than narrower baseline zones: staphylococcal ceftriaxone +1.98 mm per month (p = 0.0004) and staphylococcal cefixime +0.83 mm per month (p = 0.015). Neither prior antibiotic exposure (Spearman rho = -0.12, p = 0.31) nor subsequent doxyPEP adherence (rho = +0.04, p = 0.77) varied with enrolment date, so the cohort being sampled did not change materially over the enrolment period either (SBox 1 Figure, panel a).

Test 2. The two groups within the common window were well matched on doxyPEP tablets taken (median 42 versus 35.5, p = 0.93), prior antibiotic courses (2 versus 2, p = 0.67) and age (42 versus 40 years, p = 0.60). Within that window, month-6 specimens had smaller zone diameters than baseline specimens in 28 of 29 combinations, with a mean difference of -2.71 mm, and 8 combinations reached nominal significance despite only 39 participants contributing. The doxycycline differences were -7.96 mm in streptococci (p < 0.001), -7.74 mm in staphylococci (p = 0.017), -4.33 mm in commensal Neisseria (p = 0.010), -2.77 mm in Klebsiella (p = 0.28) and -2.52 mm in E. coli (p = 0.33) (SBox 1 Figure, panel b).

The two designs agree closely. Across the 29 combinations the same-window between-participant differences correlated with the within-participant paired changes at Spearman rho = 0.77 (p = 1e-06), with a mean absolute discrepancy of 0.99 mm, and the window estimates were if anything slightly larger in magnitude (mean -2.71 versus -2.33 mm). The single combination in which the window difference was not negative was streptococcal ceftriaxone (+0.00 mm), which is the one combination shown elsewhere in this programme to be driven by a shift in species composition rather than by susceptibility [ref] (SBox 1 Figure, panel c).

###### Conclusion and limitations

Two independent tests, one using only pre-treatment specimens and one holding the laboratory period fixed, give the same answer: the reduction in zone diameters at month 6 is not explained by secular drift in the measurement system, by seasonality, or by change in the population enrolling. Where a secular trend in baseline susceptibility exists at all it runs in the opposite direction, which would bias the trial's estimate towards the null rather than away from it.

Three limitations should be noted. The window comparison rests on 39 participants in two non-overlapping groups and is therefore underpowered for any individual combination; its value lies in the consistency of direction across 29 combinations and in its agreement with the paired analysis, not in any single p-value. The groups are balanced on the covariates we measured, but they are different people and residual confounding by unmeasured characteristics cannot be excluded, which is precisely the weakness the paired design avoids and the reason the two are presented together rather than one replacing the other. Finally, excluding laboratory drift and seasonality does not establish that doxyPEP caused the reduction; the trial remains single-arm, and elapsed time and unmeasured concurrent exposures are not separable from the intervention by either test presented here.

#### SBox 1 Figure

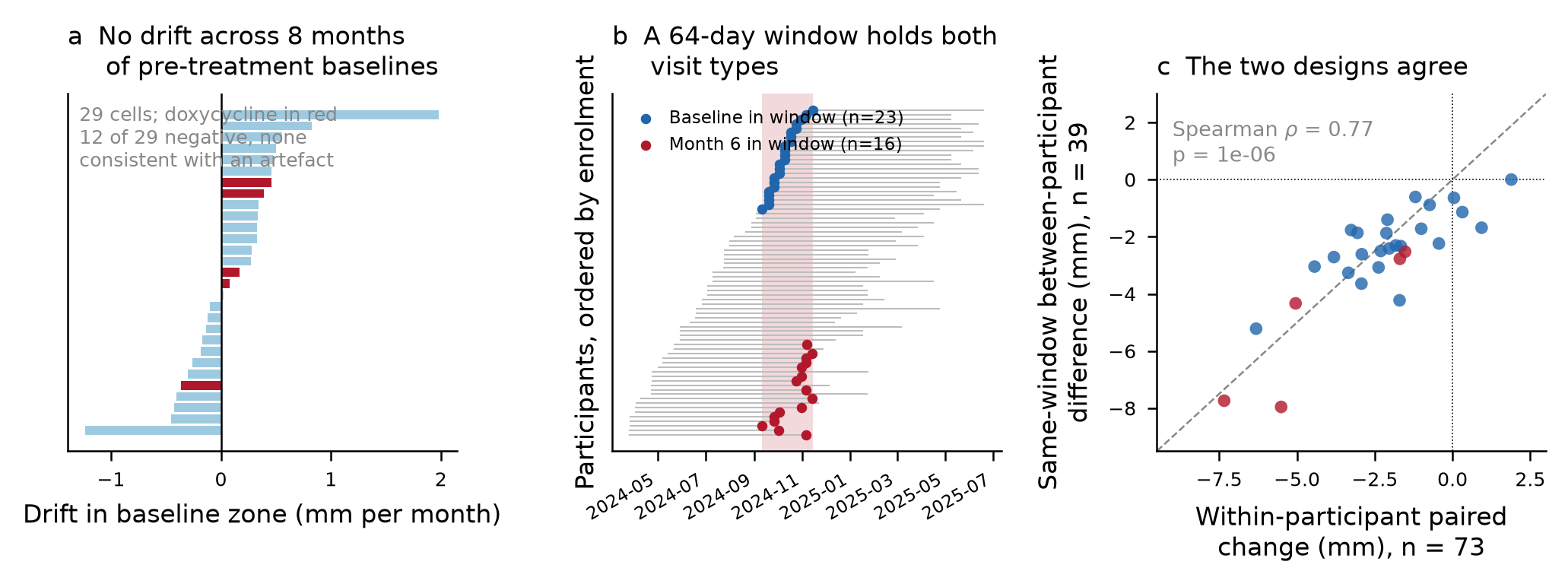

(a) Drift in baseline zone diameter with enrolment date, in millimetres per month, for each of the 29 plate-by-antimicrobial combinations; doxycycline combinations in red. All 73 specimens contributing are pre-treatment, so any slope is drift rather than effect. (b) Study visits by participant, ordered by enrolment date; grey lines join each participant's baseline and month-6 visits and the shaded band is the 64-day window in which 23 baseline and 16 month-6 visits were processed together. (c) The same-window between-participant difference against the within-participant paired change, one point per combination, doxycycline in red; dashed line is equality.

#### SBox 1 Table. Comparison within the common laboratory window

| Plate | Antimicrobial | n baseline | n month 6 | Mean difference (mm) | Mann-Whitney p | Paired change in full cohort (mm) |
| --- | --- | --- | --- | --- | --- | --- |
| Commensal Neisseria | Azithromycin | 22 | 16 | -2.41 | 0.155 | -2.03 |
| Commensal Neisseria | Cefixime | 22 | 16 | -1.40 | 0.425 | -2.09 |
| Commensal Neisseria | Ciprofloxacin | 22 | 16 | -1.86 | 0.322 | -3.06 |
| Commensal Neisseria | Ceftriaxone | 22 | 16 | -1.87 | 0.120 | -2.12 |
| Commensal Neisseria | Doxycycline | 22 | 16 | -4.33 | 0.010 | -5.04 |
| Commensal Neisseria | Penicillin | 22 | 16 | -2.33 | 0.041 | -1.66 |
| E. coli | Azithromycin | 18 | 15 | -0.89 | 0.242 | -0.74 |
| E. coli | Cefixime | 18 | 15 | -3.07 | 0.041 | -2.38 |
| E. coli | Ciprofloxacin | 18 | 15 | -2.30 | 0.759 | -1.82 |
| E. coli | Ceftriaxone | 18 | 15 | -2.50 | 0.128 | -2.31 |
| E. coli | Doxycycline | 18 | 15 | -2.52 | 0.329 | -1.52 |
| E. coli | Penicillin | 18 | 15 | -0.61 | 0.779 | -1.19 |
| Klebsiella | Azithromycin | 15 | 10 | -0.64 | 0.585 | +0.05 |
| Klebsiella | Cefixime | 15 | 10 | -3.04 | 0.617 | -4.43 |
| Klebsiella | Ciprofloxacin | 15 | 10 | -2.61 | 0.523 | -2.92 |
| Klebsiella | Ceftriaxone | 15 | 10 | -2.71 | 0.846 | -3.81 |
| Klebsiella | Doxycycline | 15 | 10 | -2.77 | 0.279 | -1.69 |
| Staphylococci | Azithromycin | 19 | 14 | -1.77 | 0.284 | -3.26 |
| Staphylococci | Cefixime | 19 | 14 | -3.64 | 0.039 | -2.93 |
| Staphylococci | Ciprofloxacin | 19 | 14 | -3.26 | 0.080 | -3.35 |
| Staphylococci | Ceftriaxone | 19 | 14 | -1.72 | 0.412 | -1.01 |
| Staphylococci | Doxycycline | 19 | 14 | -7.74 | 0.017 | -7.34 |
| Staphylococci | Penicillin | 19 | 14 | -5.22 | 0.015 | -6.31 |
| Streptococci | Azithromycin | 23 | 16 | -4.23 | 0.006 | -1.70 |
| Streptococci | Cefixime | 23 | 16 | -2.24 | 0.074 | -0.44 |
| Streptococci | Ciprofloxacin | 23 | 16 | -1.14 | 0.278 | +0.32 |
| Streptococci | Ceftriaxone | 23 | 16 | +0.00 | 0.830 | +1.89 |
| Streptococci | Doxycycline | 23 | 16 | -7.95 | 0.000 | -5.51 |
| Streptococci | Penicillin | 23 | 16 | -1.68 | 0.297 | +0.93 |

Negative differences indicate smaller zones in month-6 specimens. Klebsiella with penicillin is omitted throughout because 99.8 per cent of those zones sit at the 6 mm disc diameter.

#### Supplementary Box 2: Comparison of inhibition zone diameter against minimum inhibitory concentration in commensal *Escherichia coli*

###### Rationale

The outcome measured throughout this study is an inhibition zone diameter in millimetres. Zone diameters are a convenient and reproducible quantity, but they are not the scale in which antimicrobial resistance is conventionally expressed, and a reader presented with a change of a few millimetres has no direct way to judge how much the underlying resistance phenotype has moved. Converting millimetres into minimum inhibitory concentrations (MICs) normally requires borrowing a calibration from the literature, which is species- and method-specific and therefore a source of error. A subset of the baseline isolates in this study was tested by both methods, so the two scales can be related within the dataset itself rather than by assumption.

###### Methods

Eighty-six colonies picked from the chromogenic Escherichia coli medium at baseline, from 16 participants, had MICs determined for doxycycline, ceftriaxone and ciprofloxacin in addition to disc diffusion. Of these, 84 were identified as E. coli, one as Klebsiella oxytoca and one as Pantoea septica. No month-6 isolates and no isolates from the other four selective media were tested by both methods, so the calibration is confined to baseline E. coli.

For each antimicrobial, log2(MIC) was regressed on zone diameter by ordinary least squares, and the reciprocal of the slope taken as the zone difference corresponding to a twofold change in MIC. Observations at an assay limit were retained in the primary fit and identified separately: 12 doxycycline and 3 ciprofloxacin zones sat at the 6 mm disc diameter, and 2 doxycycline, 11 ceftriaxone and 3 ciprofloxacin MICs sat at the highest concentration tested. The doxycycline regression was repeated excluding every observation at either limit.

###### Results

Zone diameter and log2(MIC) were closely and linearly related (SFigure for SBox 2). For doxycycline the slope was -0.320 log2 units per millimetre (95% CI -0.350 to -0.289; r = -0.92, R-squared = 0.84), so 3.13 mm of zone diameter corresponds to a twofold change in MIC. Excluding all observations at an assay limit gave a nearly identical 3.05 mm per doubling (n = 73, R-squared = 0.82), so the relationship is not an artefact of the censored extremes. Ceftriaxone was steeper, at 1.91 mm per doubling (R-squared = 0.89), and ciprofloxacin shallower and considerably noisier, at 3.47 mm per doubling (R-squared = 0.57).

Applying the calibration to the six-month changes observed on the E. coli plate, the only plate for which it is valid, the doxycycline change of -1.77 mm corresponds to a 1.5-fold increase in MIC (95% CI 0.9 to 2.4), the ceftriaxone change of -1.61 mm to 1.8-fold (0.8 to 4.0), and the ciprofloxacin change of -3.03 mm to 1.8-fold (1.1 to 3.1). Expressed in the units of the composite indicator, one baseline standard deviation of the E. coli doxycycline zone distribution (6.7 mm) is equivalent to roughly a 4.4-fold change in MIC.

###### Interpretation and limitations

Three caveats bound how far this calibration can be carried. It is derived from E. coli alone, and the relationship between zone diameter and MIC depends on the organism's growth rate and on the diffusion characteristics of the agent, so the slopes reported here should not be applied to the streptococcal, staphylococcal, Neisseria or Klebsiella plates. It rests on 16 participants sampled at baseline only, which is a convenience subset rather than a random sample of the cohort. And the ceftriaxone relationship, although it has the highest R-squared of the three, is fitted across a conspicuous gap between roughly 15 and 25 mm, with a small group of high-MIC isolates anchoring one end; a slope estimated across a bimodal distribution describes the two groups' separation more than it describes a within-group gradient, and the apparent precision should be discounted accordingly.

Within those limits the calibration supports two statements that are otherwise unavailable. First, the millimetre scale used throughout this study is a close linear proxy for log2(MIC) in this organism, with doxycycline zone diameter explaining 84 per cent of the variance in doxycycline MIC; treating zone diameters as a quantitative susceptibility measure rather than dichotomising them against a breakpoint is therefore defensible on internal evidence. Second, the effect sizes reported in millimetres are modest when expressed as MIC fold-changes on this plate, in the one-and-a-half to twofold range, which is a useful corrective to reading a several-millimetre shift as a large change in phenotype.

#### SBox 2 Table. Calibration of zone diameter against MIC in baseline E. coli

| Antimicrobial | n | Slope, log2(MIC) per mm (95% CI) | r | R-squared | mm per doubling of MIC | Observations at an assay limit |
| --- | --- | --- | --- | --- | --- | --- |
| Doxycycline | 85 | -0.320 (-0.350 to -0.289) | -0.92 | 0.84 | 3.13 | 12 zones at 6 mm, 2 MICs at the assay maximum |
| Ceftriaxone | 85 | -0.525 (-0.565 to -0.485) | -0.94 | 0.89 | 1.91 | 0 zones at 6 mm, 11 MICs at the assay maximum |
| Ciprofloxacin | 85 | -0.288 (-0.343 to -0.233) | -0.75 | 0.57 | 3.47 | 3 zones at 6 mm, 3 MICs at the assay maximum |

#### SFigure for SBox 2

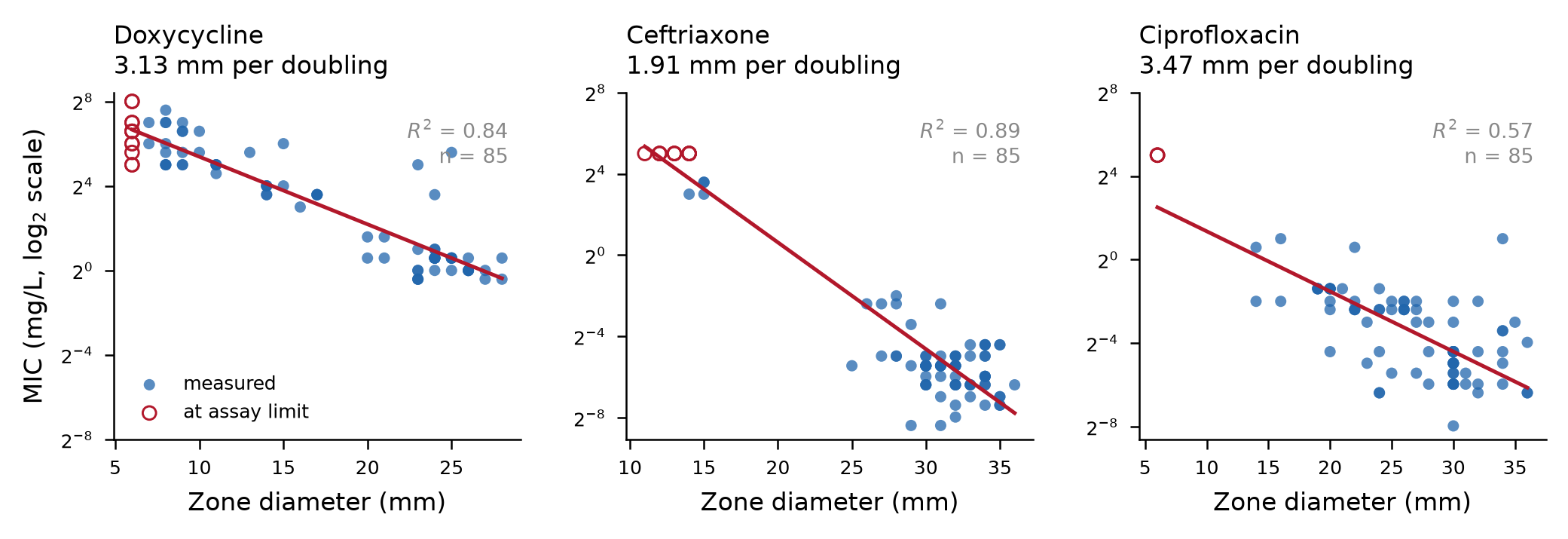

Minimum inhibitory concentration against inhibition zone diameter for 85 baseline colonies from the chromogenic E. coli medium, by antimicrobial. Red line is the ordinary least squares fit of log2(MIC) on zone diameter; open red circles are observations sitting at an assay limit (a 6 mm zone or the highest MIC tested) and were retained in the fit. The conversion quoted above each panel is the reciprocal of the fitted slope.

#### Supplementary Table S1. Sensitivity analyses for rectal inhibition-zone outcomes where 6 mm inhibition was replaced with 1 mm

| Model | LOD 1 difference (95% CrI) | LOD 1 random-slope SD (IQR) |
| --- | --- | --- |
| Doxycycline, E. coli | -1.50 (-3.90 to 0.88) | 8.19 (6.62 to 10.14) |
| Doxycycline, Klebsiella spp. | -1.05 (-3.25 to 1.33) | 7.07 (5.63 to 8.97) |
| Ceftriaxone, E. coli | -2.23 (-4.42 to 0.02) | 8.20 (6.73 to 10.02) |
| Ceftriaxone, Klebsiella spp. | -2.85 (-6.79 to 1.37) | 13.00 (10.29 to 16.65) |
| Ciprofloxacin, E. coli | -2.09 (-4.45 to 0.25) | 8.68 (7.08 to 10.65) |
| Ciprofloxacin, Klebsiella spp. | -3.83 (-8.03 to 0.23) | 12.54 (9.94 to 15.82) |
| Azithromycin, E. coli | -1.12 (-2.65 to 0.38) | 5.42 (4.42 to 6.64) |
| Azithromycin, Klebsiella spp. | -0.45 (-2.26 to 1.28) | 5.40 (4.28 to 6.80) |
| Cefixime, E. coli | -2.80 (-5.20 to -0.28) | 8.68 (7.08 to 10.62) |
| Cefixime, Klebsiella spp. | -4.54 (-8.49 to -0.79) | 12.25 (9.52 to 15.78) |

#### Supplementary Table S2. Sensitivity analyses for oral inhibition-zone outcomes where 6 mm inhibition was replaced with 1 mm

Across oral outcomes, sensitivity analyses were closely aligned with the primary findings, particularly for doxycycline and several non-tetracycline endpoints in staphylococci and commensal *Neisseria* spp.

| Model | LOD 1 difference (95% CrI) | LOD 1 random-slope SD (IQR) |
| --- | --- | --- |
| Doxycycline, oral streptococci | -5.49 (-6.81 to -4.17) | 4.43 (3.28 to 5.70) |
| Doxycycline, staphylococci | -6.68 (-9.69 to -3.53) | 10.66 (8.07 to 13.75) |
| Doxycycline, commensal Neisseria | -5.36 (-6.79 to -3.92) | 5.20 (4.08 to 6.49) |
| Ceftriaxone, oral streptococci | 1.90 (0.89 to 2.99) | 2.75 (1.17 to 4.08) |
| Ceftriaxone, staphylococci | 0.11 (-3.22 to 3.52) | 11.33 (8.63 to 14.45) |
| Ceftriaxone, commensal Neisseria | -2.16 (-3.58 to -0.64) | 5.10 (3.92 to 6.44) |
| Ciprofloxacin, oral streptococci | 0.20 (-0.85 to 1.26) | 3.11 (1.89 to 4.37) |
| Ciprofloxacin, staphylococci | -2.23 (-4.29 to -0.27) | 4.55 (1.69 to 7.22) |
| Ciprofloxacin, commensal Neisseria | -2.94 (-4.46 to -1.45) | 5.15 (3.85 to 6.64) |
| Azithromycin, oral streptococci | -2.09 (-3.47 to -0.71) | 3.36 (0.98 to 5.12) |
| Azithromycin, staphylococci | -4.29 (-7.13 to -1.68) | 8.60 (6.43 to 11.03) |
| Azithromycin, commensal Neisseria | -2.47 (-4.41 to -0.63) | 7.20 (5.76 to 8.85) |
| Cefixime, oral streptococci | -0.47 (-1.62 to 0.63) | 2.40 (0.33 to 4.10) |
| Cefixime, staphylococci | -2.70 (-4.95 to -0.17) | 7.67 (5.74 to 9.84) |
| Cefixime, commensal Neisseria | -2.12 (-3.65 to -0.60) | 5.23 (3.99 to 6.64) |
| Penicillin, oral streptococci | 0.93 (-0.45 to 2.26) | 4.02 (2.44 to 5.56) |
| Penicillin, staphylococci | -4.90 (-8.04 to -1.74) | 9.36 (6.72 to 12.39) |
| Penicillin, commensal Neisseria | -1.88 (-3.26 to -0.42) | 5.02 (3.90 to 6.35) |

#### Supplementary Table S3. Intention-to-treat analyses adjusted for cumulative doxycycline tablet use: rectal outcomes

| Model | Visit day 180 difference (95% CrI) | Tablets coefficient (95% CrI) | Random-slope SD |
| --- | --- | --- | --- |
| Doxycycline, E. coli | -1.58 (-3.79 to 0.68) | -0.02 (-0.08 to 0.05) | 8.14 |
| Doxycycline, Klebsiella spp. | -1.21 (-3.43 to 0.96) | 0.00 (-0.08 to 0.09) | 7.08 |
| Ceftriaxone, E. coli | -2.30 (-4.60 to -0.05) | -0.02 (-0.08 to 0.05) | 8.16 |
| Ceftriaxone, Klebsiella spp. | -2.85 (-6.76 to 1.10) | -0.03 (-0.14 to 0.08) | 12.98 |
| Ciprofloxacin, E. coli | -2.08 (-4.40 to 0.17) | -0.04 (-0.10 to 0.02) | 8.50 |
| Ciprofloxacin, Klebsiella spp. | -3.98 (-8.06 to 0.21) | 0.01 (-0.11 to 0.13) | 13.81 |
| Azithromycin, E. coli | -1.40 (-3.35 to 0.40) | -0.04 (-0.10 to 0.01) | 6.04 |
| Azithromycin, Klebsiella spp. | -1.90 (-4.91 to 1.17) | -0.02 (-0.11 to 0.08) | 7.30 |
| Cefixime, E. coli | -2.68 (-5.10 to -0.16) | 0.03 (-0.04 to 0.10) | 8.69 |
| Cefixime, Klebsiella spp. | -5.42 (-10.01 to -0.98) | 0.03 (-0.10 to 0.16) | 14.10 |

#### Supplementary Table S4. Intention-to-treat analyses adjusted for cumulative doxycycline tablet use: oral outcomes

| Model | Visit day 180 difference (95% CrI) | Tablets coefficient (95% CrI) | Random-slope SD |
| --- | --- | --- | --- |
| Doxycycline, oral streptococci | -5.48 (-6.75 to -4.17) | -0.05 (-0.09 to -0.01) | 4.44 |
| Doxycycline, staphylococci | -6.78 (-9.83 to -3.74) | -0.08 (-0.15 to -0.01) | 10.68 |
| Doxycycline, commensal Neisseria | -5.34 (-6.72 to -3.91) | -0.08 (-0.13 to -0.04) | 5.11 |
| Ceftriaxone, oral streptococci | 1.89 (0.82 to 2.91) | -0.03 (-0.07 to 0.00) | 2.74 |
| Ceftriaxone, staphylococci | 0.22 (-3.40 to 3.53) | -0.04 (-0.12 to 0.05) | 11.93 |
| Ceftriaxone, commensal Neisseria | -2.13 (-3.53 to -0.70) | -0.06 (-0.13 to -0.01) | 4.98 |
| Ciprofloxacin, oral streptococci | 0.25 (-0.74 to 1.27) | -0.03 (-0.06 to 0.00) | 2.86 |
| Ciprofloxacin, staphylococci | -2.31 (-4.17 to -0.34) | -0.05 (-0.14 to 0.03) | 4.55 |
| Ciprofloxacin, commensal Neisseria | -2.92 (-4.45 to -1.40) | -0.07 (-0.14 to -0.01) | 5.11 |

#### Supplementary Table S5. Per-protocol analyses for rectal outcomes

| Model | Difference (95% CrI) | Random-slope SD (IQR) |
| --- | --- | --- |
| Doxycycline, E. coli | -3.23 (-6.31 to -0.04) | 8.43 (6.41 to 11.01) |
| Doxycycline, Klebsiella spp. | -2.02 (-4.43 to 0.34) | 5.09 (3.62 to 7.20) |
| Ceftriaxone, E. coli | -2.59 (-5.75 to 0.69) | 8.61 (6.67 to 11.14) |
| Ceftriaxone, Klebsiella spp. | -1.27 (-6.50 to 4.03) | 12.90 (9.71 to 17.43) |
| Ciprofloxacin, E. coli | -2.40 (-5.70 to 0.94) | 8.92 (6.82 to 11.78) |
| Ciprofloxacin, Klebsiella spp. | -2.80 (-9.15 to 3.69) | 15.43 (11.15 to 21.34) |
| Azithromycin, E. coli | -1.46 (-3.88 to 0.80) | 5.49 (3.88 to 7.74) |
| Azithromycin, Klebsiella spp. | 0.47 (-3.46 to 4.34) | 7.05 (4.45 to 11.00) |
| Cefixime, E. coli | -2.78 (-6.22 to 0.69) | 9.43 (7.22 to 12.48) |
| Cefixime, Klebsiella spp. | -4.15 (-10.17 to 1.57) | 14.03 (9.91 to 19.97) |

#### Supplementary Table S6. Per-protocol analyses for oral outcomes

| Model | Difference (95% CrI) | Random-slope SD (IQR) |
| --- | --- | --- |
| Doxycycline, oral streptococci | -5.15 (-7.01 to -3.29) | 3.57 (1.63 to 5.55) |
| Doxycycline, staphylococci | -7.65 (-12.87 to -2.13) | 11.63 (7.39 to 17.12) |
| Doxycycline, commensal Neisseria | -6.10 (-7.93 to -4.26) | 4.21 (2.71 to 6.00) |
| Ceftriaxone, oral streptococci | 2.35 (0.48 to 4.21) | 3.40 (0.74 to 5.55) |
| Ceftriaxone, staphylococci | -2.79 (-7.67 to 1.54) | 8.77 (5.72 to 12.92) |
| Ceftriaxone, commensal Neisseria | -2.31 (-4.20 to -0.33) | 4.75 (3.34 to 6.43) |
| Ciprofloxacin, oral streptococci | -0.20 (-1.53 to 1.14) | 1.54 (0.09 to 3.30) |
| Ciprofloxacin, staphylococci | -5.33 (-7.80 to -2.72) | 3.96 (1.36 to 7.10) |
| Ciprofloxacin, commensal Neisseria | -4.54 (-6.33 to -2.42) | 4.41 (2.80 to 6.42) |
| Azithromycin, oral streptococci | -5.24 (-9.08 to -1.45) | 3.05 (0.10 to 8.63) |
| Azithromycin, staphylococci | -13.99 (-20.84 to -7.56) | 4.60 (0.14 to 14.08) |
| Azithromycin, commensal Neisseria | -3.35 (-7.43 to 0.54) | 10.14 (7.22 to 13.92) |
| Cefixime, oral streptococci | -0.35 (-2.21 to 1.56) | 2.70 (0.22 to 5.33) |
| Cefixime, staphylococci | -5.42 (-9.83 to -1.29) | 8.46 (5.63 to 12.14) |
| Cefixime, commensal Neisseria | -2.22 (-4.61 to -0.01) | 5.58 (3.88 to 7.72) |
| Penicillin, oral streptococci | 1.34 (-0.83 to 3.49) | 4.25 (1.70 to 6.79) |
| Penicillin, staphylococci | -8.56 (-13.49 to -4.03) | 7.40 (3.20 to 11.96) |
| Penicillin, commensal Neisseria | -2.21 (-4.21 to -0.20) | 4.78 (3.28 to 6.62) |

#### Supplementary Table S7. Effect of DoxyPEP in the inhibition levels among rectal samples controlling for the cumulative number of doxycycline tablets

| Model | Predictor | Difference (95% CrI) | Random slope std. deviation |
| --- | --- | --- | --- |
| DOX \| E. coli | Visit (D180) | -1.58 (-3.79 - 0.68) | 8.14 (6.63 - 10.08) |
| DOX \| E. coli | Total number of tablets | -0.02 (-0.08 - 0.05) |  |
| DOX \| Klebsiella | Visit (D180) | -1.21 (-3.43 - 0.96) | 7.08 (5.6 - 8.97) |
| DOX \| Klebsiella | Total number of tablets | 0 (-0.08 - 0.09) |  |
| CRO \| E. coli | Visit (D180) | -2.3 (-4.6 - -0.05) | 8.16 (6.7 - 9.98) |
| CRO \| E. coli | Total number of tablets | -0.02 (-0.08 - 0.05) |  |
| CRO \| Klebsiella | Visit (D180) | -2.85 (-6.76 - 1.1) | 12.98 (10.24 - 16.44) |
| CRO \| Klebsiella | Total number of tablets | -0.03 (-0.14 - 0.08) |  |
| CIP \| E. coli | Visit (D180) | -2.08 (-4.4 - 0.17) | 8.5 (6.93 - 10.46) |
| CIP \| E. coli | Total number of tablets | -0.04 (-0.1 - 0.02) |  |
| CIP \| Klebsiella | Visit (D180) | -3.98 (-8.06 - 0.21) | 13.81 (10.76 - 17.56) |
| CIP \| Klebsiella | Total number of tablets | 0.01 (-0.11 - 0.13) |  |
| AZM \| E. coli | Visit (D180) | -1.4 (-3.35 - 0.4) | 6.04 (4.7 - 7.75) |
| AZM \| E. coli | Total number of tablets | -0.04 (-0.1 - 0.01) |  |
| AZM \| Klebsiella | Visit (D180) | -1.9 (-4.91 - 1.17) | 7.3 (5.04 - 10.51) |
| AZM \| Klebsiella | Total number of tablets | -0.02 (-0.11 - 0.08) |  |
| CFM \| E. coli | Visit (D180) | -2.68 (-5.1 - -0.16) | 8.69 (7.06 - 10.75) |
| CFM \| E. coli | Total number of tablets | 0.03 (-0.04 - 0.1) |  |
| CFM \| Klebsiella | Visit (D180) | -5.42 (-10.01 - -0.98) | 14.1 (10.71 - 18.63) |
| CFM \| Klebsiella | Total number of tablets | 0.03 (-0.1 - 0.16) |  |

#### Supplementary Table S8. Effect of DoxyPEP in the inhibition levels among oral samples controlling for the cumulative number of doxycycline tablets

| Model | Predictor | Difference (95% CrI) | Random slope std. deviation |
| --- | --- | --- | --- |
| DOX \| Oral streptococci | Visit (D180) | -5.48 (-6.75 - -4.17) | 4.44 (3.33 - 5.67) |
| DOX \| Oral streptococci | Total number of tablets | -0.05 (-0.09 - -0.01) |  |
| DOX \| Staphylococcus aureus | Visit (D180) | -6.78 (-9.83 - -3.74) | 10.68 (8.05 - 13.83) |
| DOX \| Staphylococcus aureus | Total number of tablets | -0.08 (-0.15 - -0.01) |  |
| DOX \| Commensal Neisseria | Visit (D180) | -5.34 (-6.72 - -3.91) | 5.11 (4 - 6.39) |
| DOX \| Commensal Neisseria | Total number of tablets | -0.08 (-0.13 - -0.04) |  |
| CRO \| Oral streptococci | Visit (D180) | 1.89 (0.82 - 2.91) | 2.74 (1.24 - 4.09) |
| CRO \| Oral streptococci | Total number of tablets | -0.03 (-0.07 - 0) |  |
| CRO \| Staphylococcus aureus | Visit (D180) | 0.22 (-3.4 - 3.53) | 11.93 (8.95 - 15.48) |
| CRO \| Staphylococcus aureus | Total number of tablets | -0.04 (-0.12 - 0.05) |  |
| CRO \| Commensal Neisseria | Visit (D180) | -2.13 (-3.53 - -0.7) | 4.98 (3.86 - 6.26) |
| CRO \| Commensal Neisseria | Total number of tablets | -0.06 (-0.13 - -0.01) |  |
| CIP \| Oral streptococci | Visit (D180) | 0.25 (-0.74 - 1.27) | 2.86 (1.7 - 3.99) |
| CIP \| Oral streptococci | Total number of tablets | -0.03 (-0.06 - 0) |  |
| CIP \| Staphylococcus aureus | Visit (D180) | -2.31 (-4.17 - -0.34) | 4.55 (1.82 - 7.13) |
| CIP \| Staphylococcus aureus | Total number of tablets | -0.05 (-0.14 - 0.03) |  |
| CIP \| Commensal Neisseria | Visit (D180) | -2.92 (-4.45 - -1.4) | 5.11 (3.8 - 6.63) |
| CIP \| Commensal Neisseria | Total number of tablets | -0.07 (-0.14 - -0.01) |  |
| AZM \| Oral streptococci | Visit (D180) | -4.49 (-7.3 - -1.79) | 5.76 (0.68 - 10.08) |
| AZM \| Oral streptococci | Total number of tablets | -0.14 (-0.25 - -0.04) |  |
| AZM \| Staphylococcus aureus | Visit (D180) | -9.33 (-14.63 - -4.28) | 15.43 (10.61 - 21.16) |
| AZM \| Staphylococcus aureus | Total number of tablets | -0.18 (-0.34 - -0.04) |  |
| AZM \| Commensal Neisseria | Visit (D180) | -3.12 (-5.77 - -0.67) | 9.32 (7.32 - 11.79) |
| AZM \| Commensal Neisseria | Total number of tablets | -0.13 (-0.22 - -0.05) |  |
| CFM \| Oral streptococci | Visit (D180) | -0.48 (-1.62 - 0.6) | 2.39 (0.41 - 4.08) |
| CFM \| Oral streptococci | Total number of tablets | -0.05 (-0.09 - -0.01) |  |
| CFM \| Staphylococcus aureus | Visit (D180) | -3.1 (-5.34 - -0.5) | 7.91 (5.94 - 10.28) |
| CFM \| Staphylococcus aureus | Total number of tablets | -0.1 (-0.18 - -0.03) |  |
| CFM \| Commensal Neisseria | Visit (D180) | -2.05 (-3.58 - -0.62) | 5.22 (3.99 - 6.6) |
| CFM \| Commensal Neisseria | Total number of tablets | -0.09 (-0.16 - -0.01) |  |
| PEN \| Oral streptococci | Visit (D180) | 0.91 (-0.49 - 2.25) | 4.01 (2.41 - 5.57) |
| PEN \| Oral streptococci | Total number of tablets | -0.04 (-0.07 - 0) |  |
| PEN \| Staphylococcus aureus | Visit (D180) | -4.87 (-8.2 - -1.64) | 9.57 (6.78 - 12.75) |
| PEN \| Staphylococcus aureus | Total number of tablets | -0.07 (-0.15 - 0.01) |  |
| PEN \| Commensal Neisseria | Visit (D180) | -1.78 (-3.13 - -0.47) | 4.73 (3.63 - 5.94) |
| PEN \| Commensal Neisseria | Total number of tablets | -0.04 (-0.09 - 0) |  |

#### Supplementary Table S9. Effect of DoxyPEP in the inhibition levels among rectal samples controlling for other antibiotic use during the study

| Model | Predictor | Difference (95% CrI) | Random slope std. deviation |
| --- | --- | --- | --- |
| DOX \| E. coli | Visit (D180) | -1.59 (-3.77 - 0.68) | 8.14 (6.62 - 10.01) |
| DOX \| E. coli | Antibiotics during study | -1.54 (-3.93 - 0.91) |  |
| DOX \| Klebsiella | Visit (D180) | -1.22 (-3.33 - 0.98) | 7.01 (5.57 - 8.83) |
| DOX \| Klebsiella | Antibiotics during study | -0.78 (-3.63 - 2.16) |  |
| CRO \| E. coli | Visit (D180) | -2.22 (-4.54 - -0.02) | 8.11 (6.67 - 9.93) |
| CRO \| E. coli | Antibiotics during study | -1.15 (-3.35 - 1.05) |  |
| CRO \| Klebsiella | Visit (D180) | -2.8 (-6.75 - 1.55) | 13.2 (10.37 - 16.99) |
| CRO \| Klebsiella | Antibiotics during study | -1.41 (-5 - 2.15) |  |
| CIP \| E. coli | Visit (D180) | -2.09 (-4.32 - 0.25) | 8.49 (6.94 - 10.35) |
| CIP \| E. coli | Antibiotics during study | -0.93 (-2.96 - 1.26) |  |
| CIP \| Klebsiella | Visit (D180) | -3.91 (-8.17 - 0.37) | 13.68 (10.84 - 17.22) |
| CIP \| Klebsiella | Antibiotics during study | 2.18 (-2.25 - 6.07) |  |
| AZM \| E. coli | Visit (D180) | -1.35 (-3.22 - 0.41) | 5.99 (4.61 - 7.81) |
| AZM \| E. coli | Antibiotics during study | -2.46 (-4.46 - -0.39) |  |
| AZM \| Klebsiella | Visit (D180) | -1.72 (-4.76 - 1.27) | 7.14 (4.95 - 10.16) |
| AZM \| Klebsiella | Antibiotics during study | -1.99 (-5.66 - 1.09) |  |
| CFM \| E. coli | Visit (D180) | -2.76 (-5.01 - -0.41) | 8.69 (7.06 - 10.66) |
| CFM \| E. coli | Antibiotics during study | -0.49 (-2.93 - 1.93) |  |
| CFM \| Klebsiella | Visit (D180) | -5.45 (-10.04 - -0.86) | 14.24 (10.81 - 19.13) |
| CFM \| Klebsiella | Antibiotics during study | -0.66 (-5.09 - 3.73) |  |

#### Supplementary Table S10. Effect of DoxyPEP in the inhibition levels among oral samples controlling for other antibiotic use during the study

| Model | Predictor | Difference (95% CrI) | Random slope std. deviation |
| --- | --- | --- | --- |
| DOX \| Oral streptococci | Visit (D180) | -5.46 (-6.8 - -4.21) | 4.42 (3.25 - 5.7) |
| DOX \| Oral streptococci | Antibiotics during study | -0.89 (-2.4 - 0.62) |  |
| DOX \| Staphylococcus aureus | Visit (D180) | -6.72 (-9.84 - -3.62) | 10.84 (8.2 - 14.01) |
| DOX \| Staphylococcus aureus | Antibiotics during study | -1.66 (-4.13 - 0.7) |  |
| DOX \| Commensal Neisseria | Visit (D180) | -5.29 (-6.71 - -3.9) | 5.13 (3.99 - 6.38) |
| DOX \| Commensal Neisseria | Antibiotics during study | -0.3 (-2.16 - 1.46) |  |
| CRO \| Oral streptococci | Visit (D180) | 1.9 (0.81 - 2.94) | 2.74 (1.21 - 4.08) |
| CRO \| Oral streptococci | Antibiotics during study | -1.27 (-2.43 - -0.08) |  |
| CRO \| Staphylococcus aureus | Visit (D180) | 0.25 (-3.12 - 3.78) | 11.96 (9.07 - 15.43) |
| CRO \| Staphylococcus aureus | Antibiotics during study | -0.08 (-3.25 - 2.95) |  |
| CRO \| Commensal Neisseria | Visit (D180) | -2.08 (-3.49 - -0.61) | 4.97 (3.85 - 6.27) |
| CRO \| Commensal Neisseria | Antibiotics during study | -2.19 (-4.45 - 0.03) |  |
| CIP \| Oral streptococci | Visit (D180) | 0.25 (-0.77 - 1.2) | 2.86 (1.78 - 3.97) |
| CIP \| Oral streptococci | Antibiotics during study | -0.68 (-1.82 - 0.43) |  |
| CIP \| Staphylococcus aureus | Visit (D180) | -2.27 (-4.23 - -0.42) | 4.55 (2.01 - 7.11) |
| CIP \| Staphylococcus aureus | Antibiotics during study | 0.4 (-2.6 - 3.41) |  |
| CIP \| Commensal Neisseria | Visit (D180) | -2.88 (-4.32 - -1.38) | 5.1 (3.8 - 6.57) |
| CIP \| Commensal Neisseria | Antibiotics during study | -1.99 (-4.33 - 0.27) |  |
| AZM \| Oral streptococci | Visit (D180) | -4.59 (-7.46 - -1.82) | 6.31 (1.31 - 10.42) |
| AZM \| Oral streptococci | Antibiotics during study | -3.55 (-7.52 - 0.27) |  |
| AZM \| Staphylococcus aureus | Visit (D180) | -8.81 (-14.16 - -3.74) | 15.72 (10.95 - 21.55) |
| AZM \| Staphylococcus aureus | Antibiotics during study | -1.47 (-6.56 - 3.9) |  |
| AZM \| Commensal Neisseria | Visit (D180) | -3 (-5.57 - -0.59) | 9.27 (7.26 - 11.58) |
| AZM \| Commensal Neisseria | Antibiotics during study | -2.5 (-5.64 - 0.67) |  |
| CFM \| Oral streptococci | Visit (D180) | -0.49 (-1.62 - 0.64) | 2.42 (0.32 - 4.08) |
| CFM \| Oral streptococci | Antibiotics during study | -1.79 (-3.2 - -0.34) |  |
| CFM \| Staphylococcus aureus | Visit (D180) | -3 (-5.62 - -0.52) | 8.25 (6.2 - 10.74) |
| CFM \| Staphylococcus aureus | Antibiotics during study | -1.12 (-4 - 1.49) |  |
| CFM \| Commensal Neisseria | Visit (D180) | -2.04 (-3.49 - -0.5) | 5.26 (4.03 - 6.67) |
| CFM \| Commensal Neisseria | Antibiotics during study | -2.63 (-5.2 - -0.01) |  |
| PEN \| Oral streptococci | Visit (D180) | 0.92 (-0.43 - 2.28) | 3.99 (2.41 - 5.49) |
| PEN \| Oral streptococci | Antibiotics during study | -1.6 (-2.79 - -0.21) |  |
| PEN \| Staphylococcus aureus | Visit (D180) | -4.87 (-8.03 - -1.64) | 9.81 (6.95 - 12.98) |
| PEN \| Staphylococcus aureus | Antibiotics during study | -0.04 (-3.07 - 2.71) |  |
| PEN \| Commensal Neisseria | Visit (D180) | -1.77 (-3.12 - -0.46) | 4.64 (3.58 - 5.86) |
| PEN \| Commensal Neisseria | Antibiotics during study | -1.27 (-2.88 - 0.37) |  |

#### Supplementary Table S11. Effect of DoxyPEP in the inhibition levels among rectal samples for the sub-group of high doxycycline consumption

| Model | Difference (95% CrI) | Random slope std. deviation |
| --- | --- | --- |
| DOX \| E. coli | -2.42 (-5.87 - 0.93) | 7.31 (5.03 - 10.41) |
| DOX \| Klebsiella | -1.65 (-5.58 - 2.49) | 7.59 (5.15 - 11.16) |
| CRO \| E. coli | -3.84 (-8.15 - 0.45) | 9.77 (7.17 - 13.51) |
| CRO \| Klebsiella | -9.56 (-16.48 - -2.19) | 14.4 (9.73 - 21.69) |
| CIP \| E. coli | -3.3 (-7.4 - 0.41) | 8.63 (6.2 - 12.16) |
| CIP \| Klebsiella | -6.35 (-12.77 - 0.64) | 13.34 (8.78 - 20.31) |
| AZM \| E. coli | -0.8 (-3.86 - 2.5) | 6.16 (4.14 - 9.04) |
| AZM \| Klebsiella | -1.33 (-6.64 - 3.93) | 8.45 (4.9 - 14.78) |
| CFM \| E. coli | -3.97 (-7.99 - 0.35) | 9.53 (6.98 - 13.19) |
| CFM \| Klebsiella | -10.44 (-18.89 - -2.32) | 15.68 (9.85 - 25.53) |

#### Supplementary Table S12. Effect of DoxyPEP in the inhibition levels among oral samples for the sub-group of high doxycycline consumption

| Model | Difference (95% CrI) | Random slope std. deviation |
| --- | --- | --- |
| DOX \| Oral streptococci | -4.69 (-6.58 - -2.83) | 3.63 (2.04 - 5.49) |
| DOX \| Staphylococcus aureus | -2.9 (-8.1 - 2.21) | 10.95 (7.29 - 15.63) |
| DOX \| Commensal Neisseria | -3.28 (-5.71 - -0.72) | 5.64 (3.88 - 7.87) |
| CRO \| Oral streptococci | 2.02 (0.58 - 3.61) | 2 (0.15 - 4.23) |
| CRO \| Staphylococcus aureus | 1.05 (-3.18 - 5.43) | 5.37 (0.45 - 11.85) |
| CRO \| Commensal Neisseria | 0.11 (-2.09 - 2.42) | 4.68 (2.96 - 6.78) |
| CIP \| Oral streptococci | 1.15 (-0.57 - 2.84) | 2.59 (0.5 - 4.66) |
| CIP \| Staphylococcus aureus | 1.73 (-1.99 - 5.68) | 5.02 (1.21 - 9.46) |
| CIP \| Commensal Neisseria | -0.02 (-2.6 - 2.75) | 5.89 (3.78 - 8.55) |
| AZM \| Oral streptococci | -5.31 (-9.06 - -1.74) | 1.94 (0.08 - 5.82) |
| AZM \| Staphylococcus aureus | 1.6 (-5.97 - 9.44) | 10.66 (1.69 - 20.83) |
| AZM \| Commensal Neisseria | -1.08 (-5.26 - 3.23) | 8.88 (5.38 - 13.34) |
| CFM \| Oral streptococci | -0.56 (-2.55 - 1.55) | 2.94 (0.67 - 5.31) |
| CFM \| Staphylococcus aureus | -0.78 (-4.94 - 2.93) | 5.48 (0.73 - 10.76) |
| CFM \| Commensal Neisseria | 0.68 (-1.66 - 3.03) | 4.65 (2.64 - 6.91) |
| PEN \| Oral streptococci | 1.37 (-0.6 - 3.42) | 3 (0.52 - 5.53) |
| PEN \| Staphylococcus aureus | -0.42 (-4.71 - 3.78) | 4.9 (0.36 - 10.51) |
| PEN \| Commensal Neisseria | 0.45 (-2.14 - 3.17) | 5.85 (3.88 - 8.43) |

### Supplementary Table S13. Estimated change from baseline to day 180 in rectal inhibition-zone diameter, intention-to-treat analysis

| Antibiotic | Organism | Difference, mm (95% CrI) | Random-slope SD, mm (95% CrI) |
| --- | --- | --- | --- |
| Doxycycline | *E. coli* | -1.67 (-4.02 to 0.49) | 8.16 (6.64 to 10.05) |
| Cefixime | *E. coli* | -2.69 (-5.12 to -0.29) | 8.66 (7.02 to 10.65) |
| Ciprofloxacin | *E. coli* | -2.15 (-4.46 to 0.12) | 8.56 (6.93 to 10.50) |
| Ceftriaxone | E. coli | -2.17 (-4.43 to 0.15) | 8.15 (6.68 to 10.02) |
| Azithromycin | E. coli | -1.35 (-3.18 to 0.43) | 5.98 (4.60 to 7.72) |
| Doxycycline | *Klebsiella spp.* | -1.25 (-3.49 to 1.00) | 7.07 (5.60 to 8.98) |
| Cefixime | *Klebsiella spp.* | -5.44 (-9.75 to -0.90) | 14.10 (10.78 to 18.49) |
| Ciprofloxacin | *Klebsiella spp.* | -4.04 (-8.47 to 0.11) | 13.78 (10.75 to 17.73) |
| Ceftriaxone | Klebsiella spp. | -2.75 (-6.67 to 1.42) | 13.01 (10.23 to 16.63) |
| Azithromycin | Klebsiella spp. | -1.92 (-5.25 to 1.14) | 7.28 (5.05 to 10.53) |

### Supplementary Table S14.. Estimated change from baseline to day 180 in oral inhibition-zone diameter, intention-to-treat analysis

| Antibiotic | Organism | Difference, mm (95% CrI) | Random-slope SD, mm (95% CrI) |
| --- | --- | --- | --- |
| Doxycycline | Streptococci | -5.47 (-6.75 to -4.15) | 4.41 (3.29 to 5.69) |
| Cefixime | Streptococci | -0.48 (-1.60 to 0.68) | 2.41 (0.40 to 4.11) |
| Ciprofloxacin | Streptococci | 0.26 (-0.71 to 1.26) | 2.86 (1.74 to 3.97) |
| Ceftriaxone | Streptococci | 1.89 (0.81 to 2.93) | 2.71 (0.99 to 4.06) |
| Azithromycin | Streptococci | -4.56 (-7.37 to -1.68) | 6.10 (1.02 to 10.36) |
| Penicillin | Streptococci | 0.92 (-0.44 to 2.32) | 4.00 (2.46 to 5.54) |
| Doxycycline | Staphylococci | -6.66 (-10.09 to -3.67) | 10.77 (8.05 to 14.06) |
| Cefixime | Staphylococci | -2.96 (-5.46 to -0.47) | 8.27 (6.23 to 10.66) |
| Ciprofloxacin | Staphylococci | -2.29 (-4.30 to -0.38) | 4.54 (1.94 to 7.05) |
| Ceftriaxone | Staphylococci | 0.26 (-3.29 to 3.80) | 12.00 (9.17 to 15.51) |
| Azithromycin | Staphylococci | -8.76 (-14.02 to -3.39) | 15.62 (10.86 to 21.37) |
| Penicillin | Staphylococci | -4.80 (-8.10 to -1.58) | 9.77 (7.03 to 13.00) |
| Doxycycline | Commensal *Neisseria* | -5.28 (-6.62 to -3.86) | 5.12 (4.01 to 6.43) |
| Cefixime | Commensal *Neisseria* | -2.01 (-3.49 to -0.51) | 5.26 (4.09 to 6.66) |
| Ciprofloxacin | Commensal *Neisseria* | -2.90 (-4.39 to -1.40) | 5.12 (3.87 to 6.56) |
| Ceftriaxone | Commensal Neisseria | -2.05 (-3.43 to -0.60) | 4.95 (3.86 to 6.25) |
| Azithromycin | Commensal Neisseria | -3.08 (-5.63 to -0.66) | 9.31 (7.35 to 11.68) |
| Penicillin | Commensal Neisseria | -1.76 (-3.05 to -0.47) | 4.66 (3.62 to 5.88) |

### Supplementary Figure S1: Number of doxycycline PEP (200mg) doses since last visit

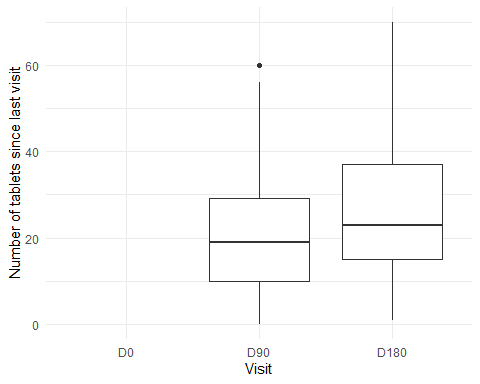

### Supplementary Figure S2: Frequency of condom use during anal sex

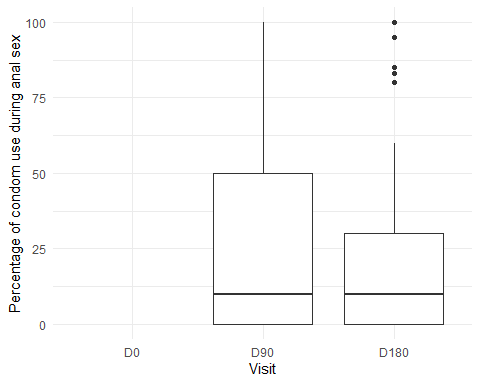

### Supplementary Figure S3: Proportion of condom use since study commencement

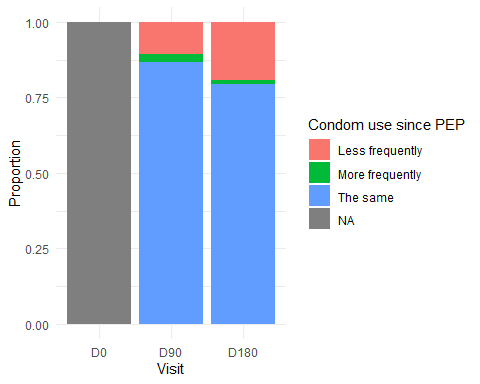

### Supplementary Figure S4: Number of reported sex partners per study visit (median [thick black line] and IQR [box]).

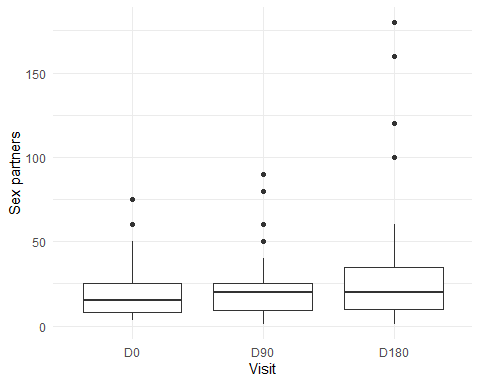

#### Supplementary Figure S5. Histograms of inhibition-zone distributions for each organism-antibiotic combination at baseline and day 180.

##### Distribution of inhibition zone diameters among rectal samples

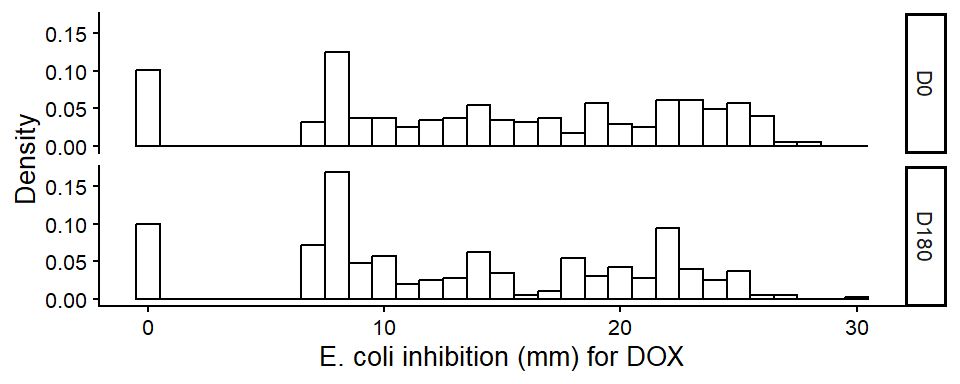

Distribution of inhibition zone for E. coli on doxycycline

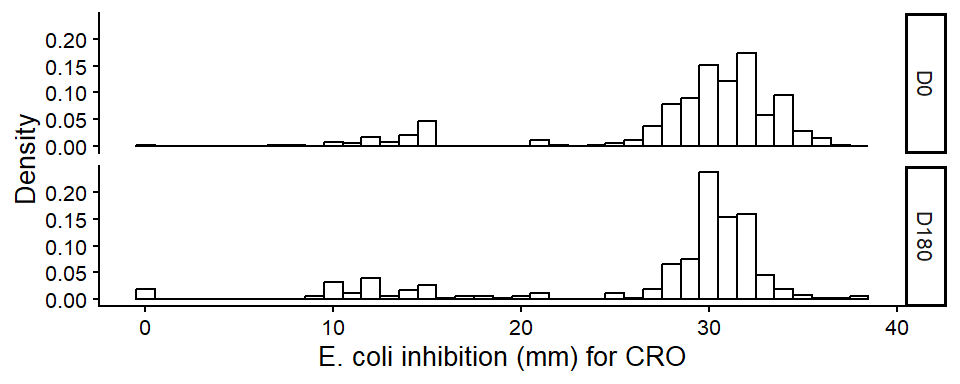

Distribution of inhibition zone for E. coli on ceftriaxone

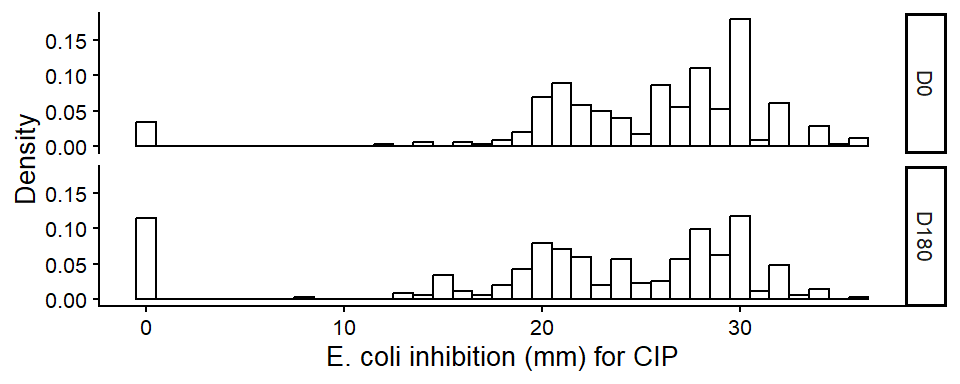

  Distribution of inhibition zone for E. coli on ciprofloxacin

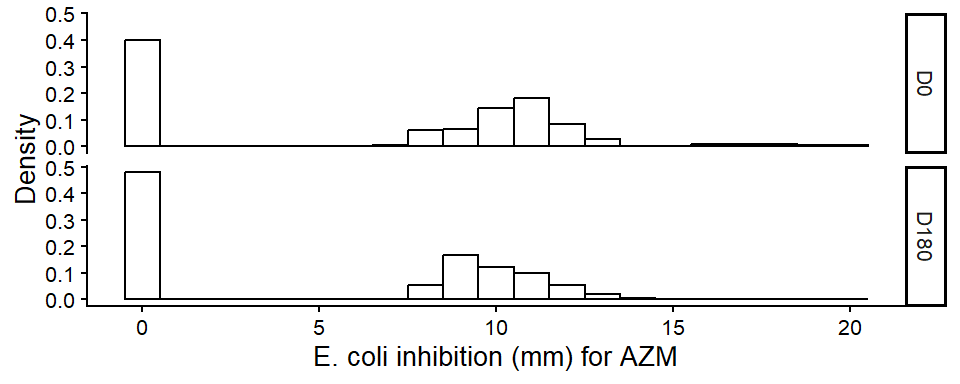

  Distribution of inhibition zone for E. coli on azithromycin

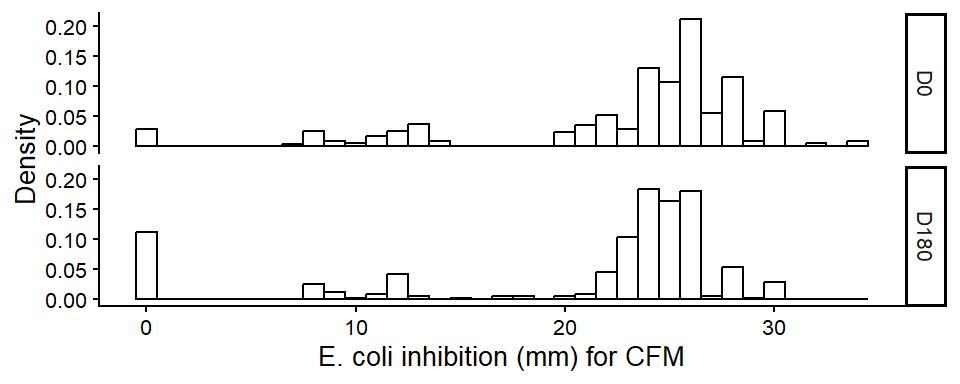

  Distribution of inhibition zone for E. coli on cefixime

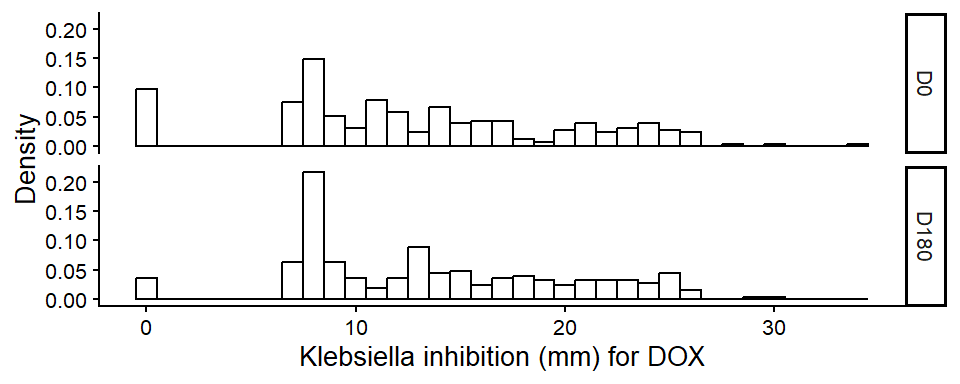

Distribution of inhibition zone for Klebsiella pneumoniae on doxycycline

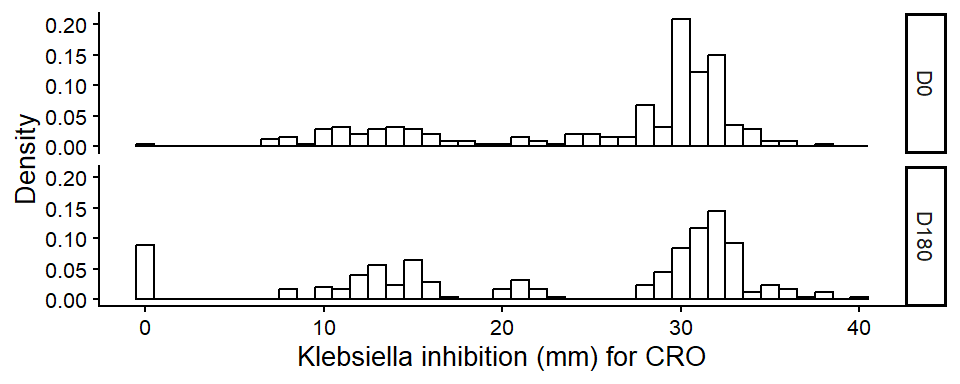

  Distribution of inhibition zone for Klebsiella pneumoniae on ceftriaxone

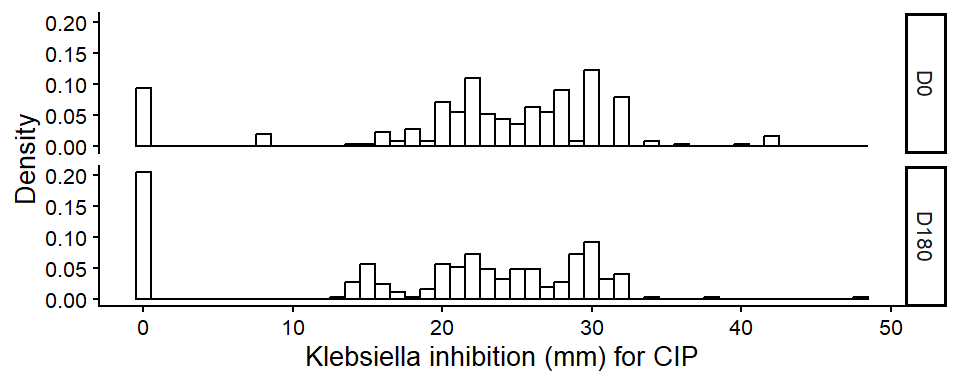

  Distribution of inhibition zone for Klebsiella pneumoniae on ciprofloxacin

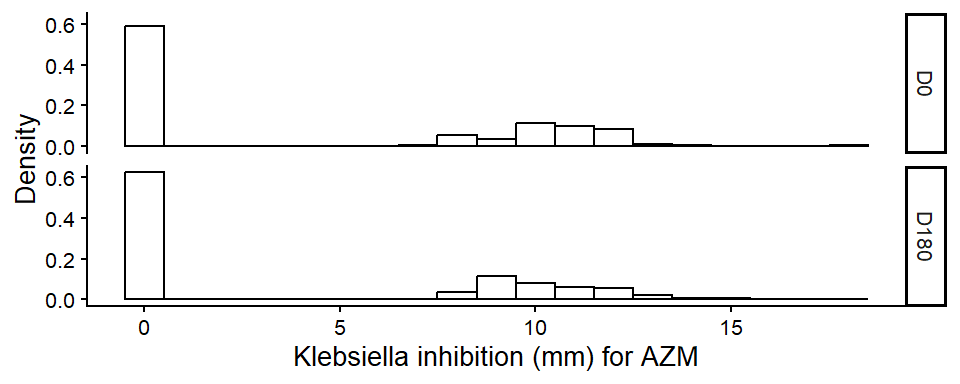

  Distribution of inhibition zone for Klebsiella pneumoniae on azithromycin

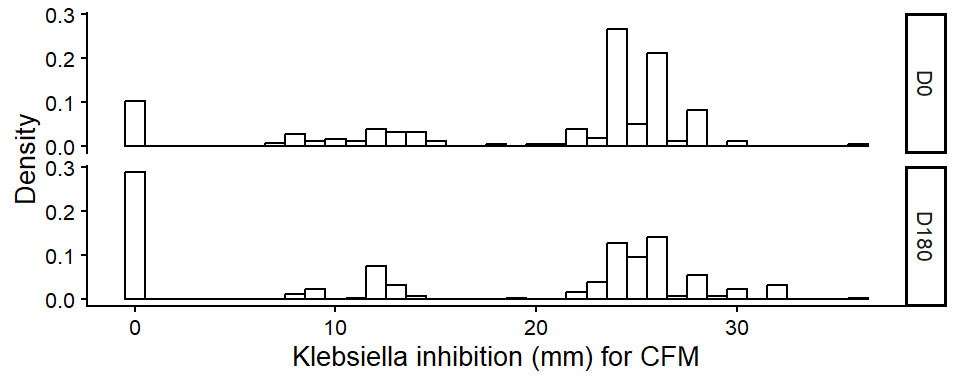

  Distribution of inhibition zone for Klebsiella pneumoniae on cefixime

##### Distribution of inhibition zone diameters among oral samples

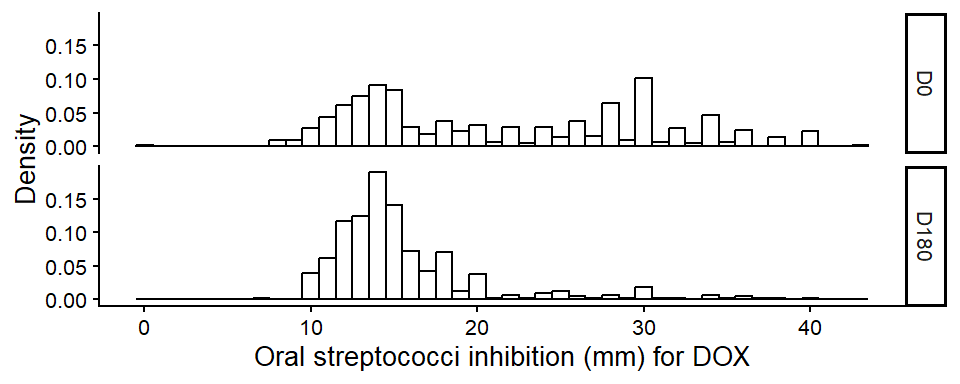

  Distribution of inhibition zone for oral streptococci on doxycycline

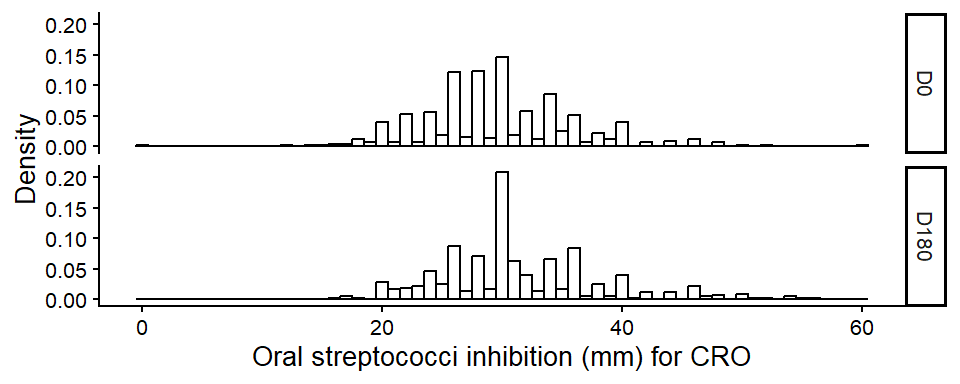

  Distribution of inhibition zone for oral streptococci on ceftriaxone

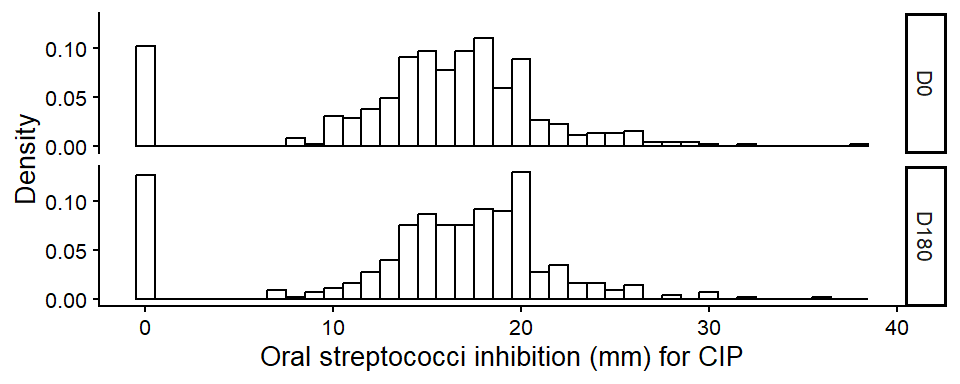

  Distribution of inhibition zone for oral streptococci on ciprofloxacin

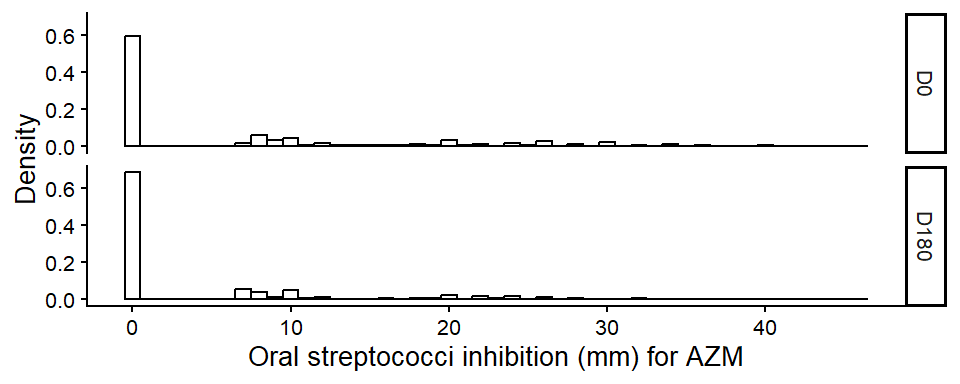

  Distribution of inhibition zone for oral streptococci on azithromycin

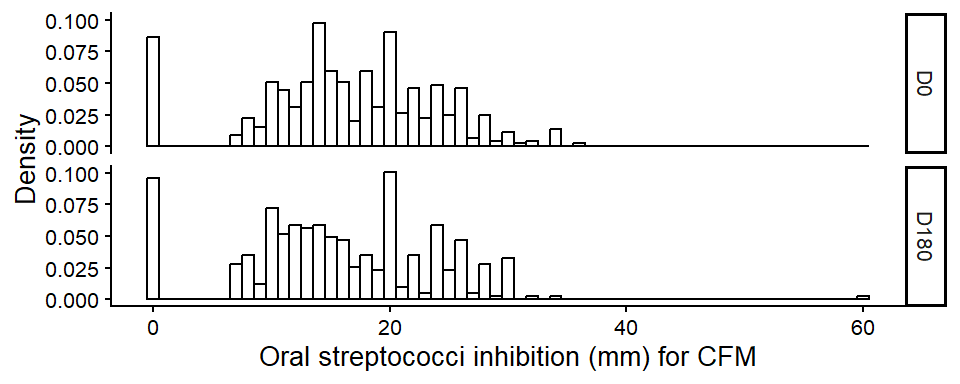

Distribution of inhibition zone for oral streptococci on cefixime

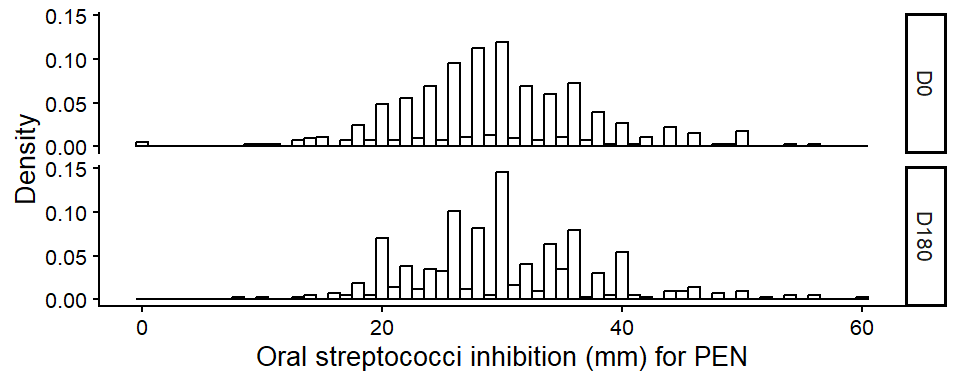

  Distribution of inhibition zone for oral streptococci on penicillin

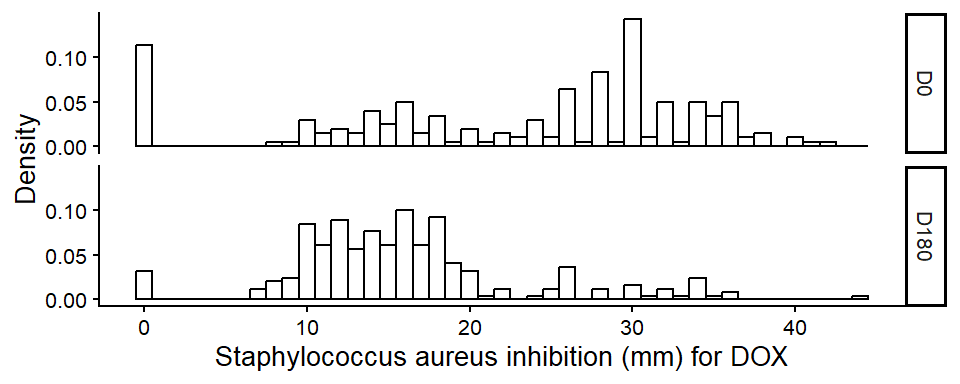

  Distribution of inhibition zone for Staphylococcus aureus on doxycycline

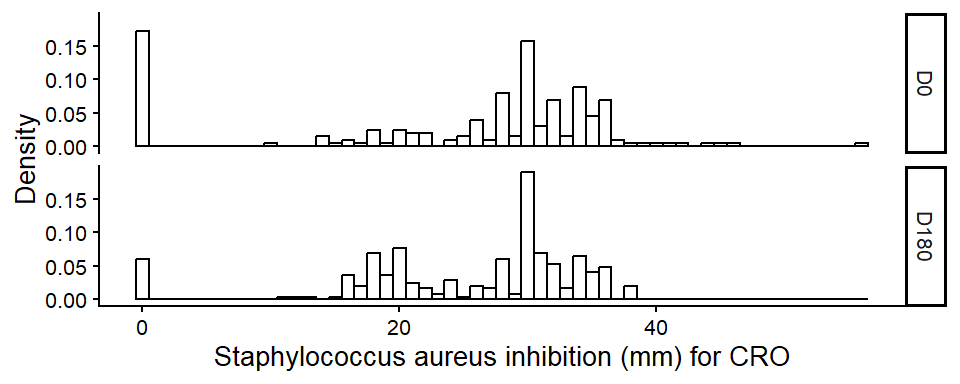

  Distribution of inhibition zone for Staphylococcus aureus on ceftriaxone

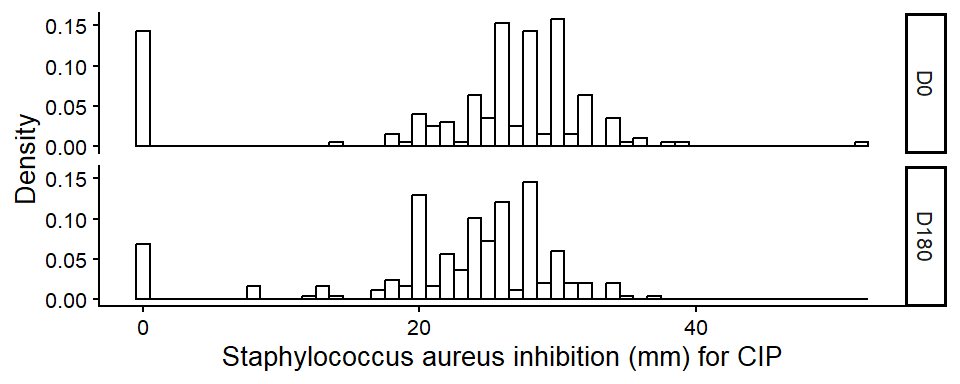

Distribution of inhibition zone for Staphylococcus aureus on ciprofloxacin

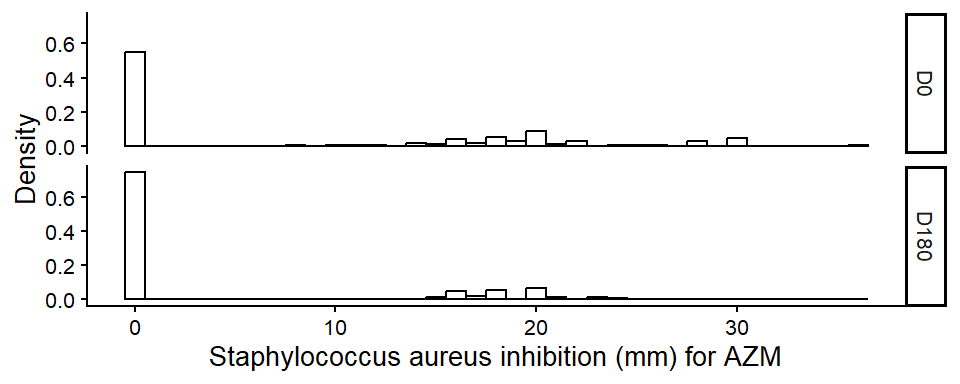

  Distribution of inhibition zone for Staphylococcus aureus on azithromycin

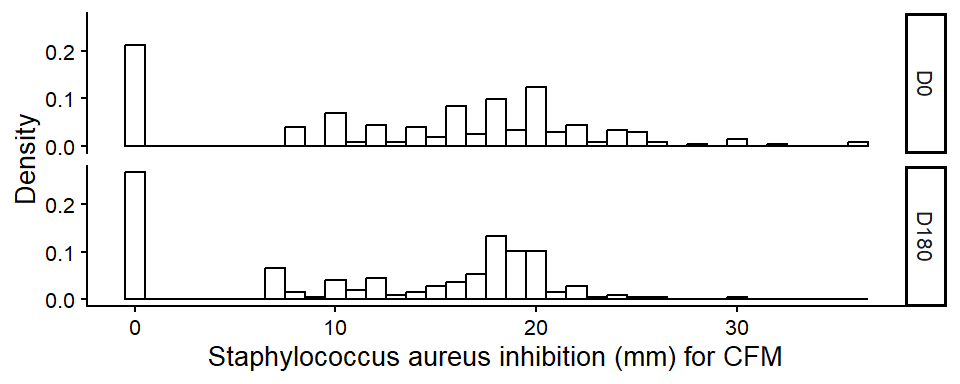

  Distribution of inhibition zone for Staphylococcus aureus on cefixime

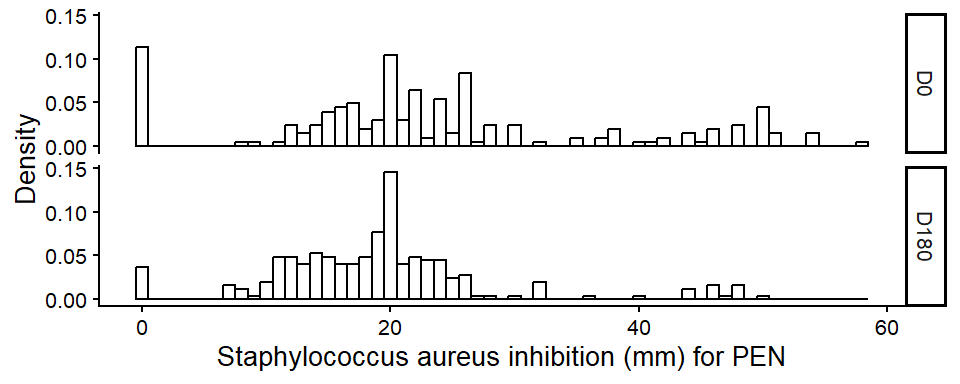

  Distribution of inhibition zone for Staphylococcus aureus on penicillin

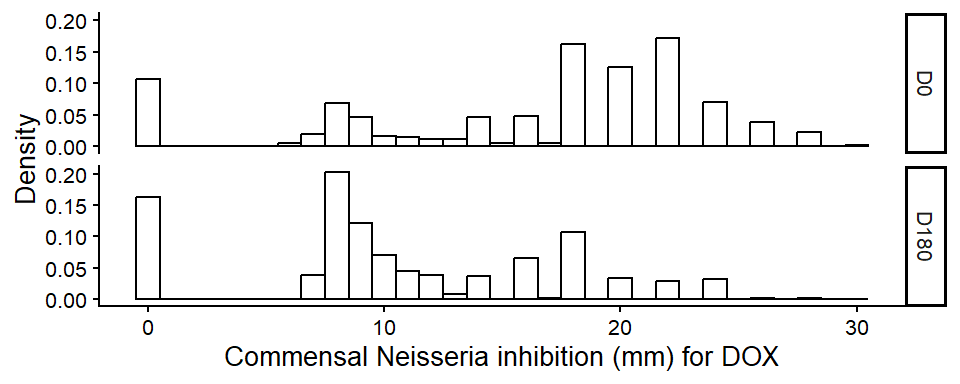

  Distribution of inhibition zone for Commensal Neisseria on doxycycline

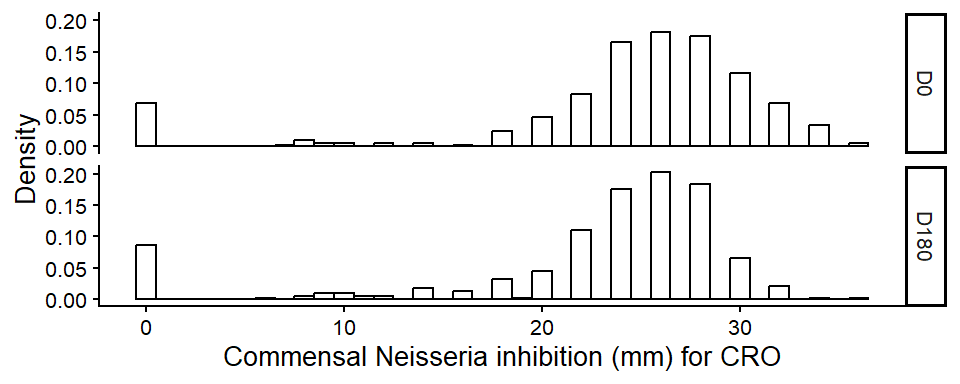

  Distribution of inhibition zone for Commensal Neisseria on ceftriaxone

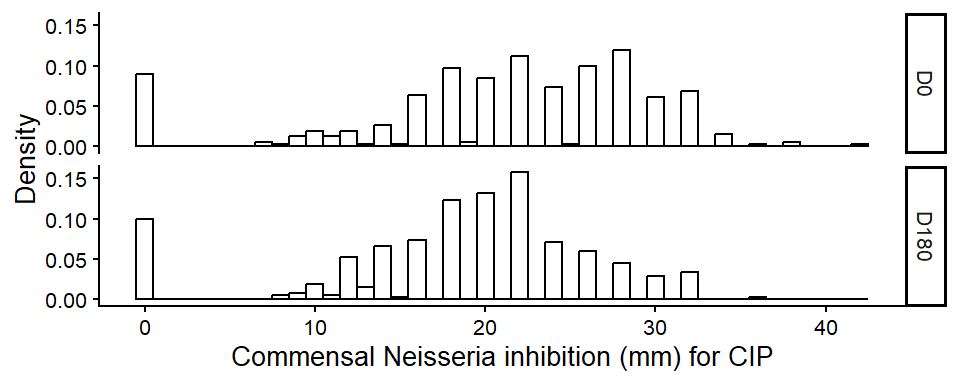

  Distribution of inhibition zone for Commensal Neisseria on ciprofloxacin

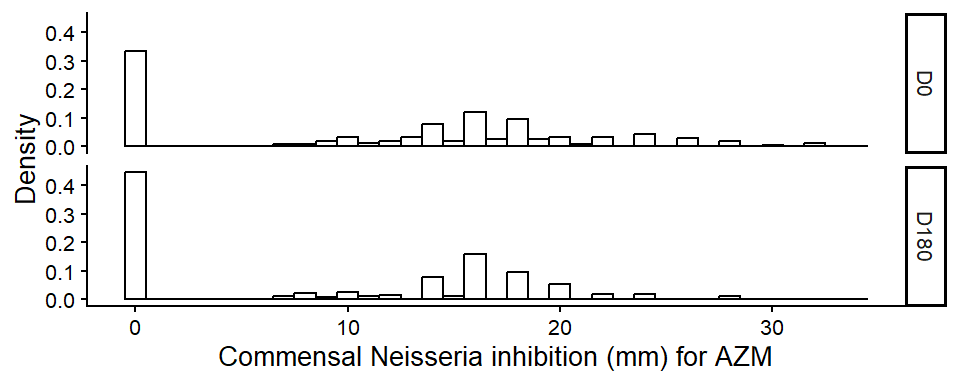

  Distribution of inhibition zone for Commensal Neisseria on azithromycin

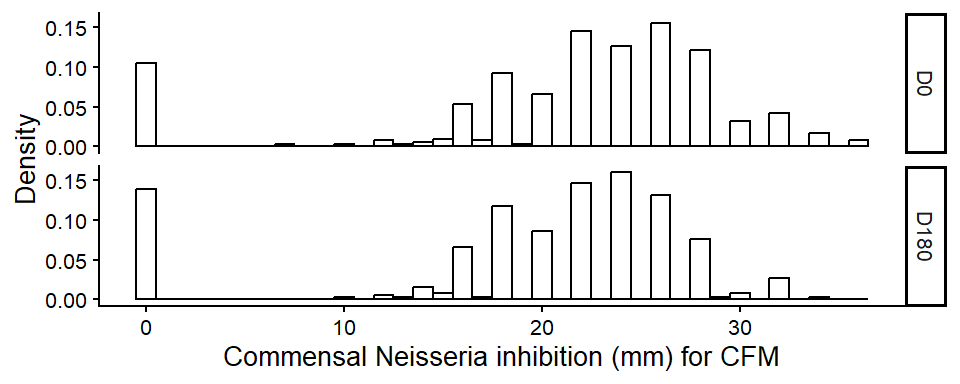

Distribution of inhibition zone for Commensal Neisseria on cefixime

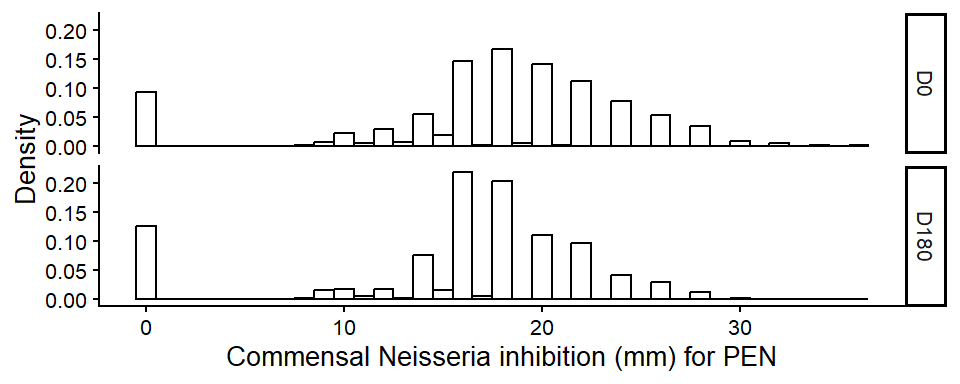

  Distribution of inhibition zone for Commensal Neisseria on penicillin

#### Supplementary figure S6. Individual shift of susceptibility among samples

The individual trajectories in terms of inhibition were plotted for each combination of bacteria and antibiotics in the study. All participants in the intention to treat population were included in this analysis.

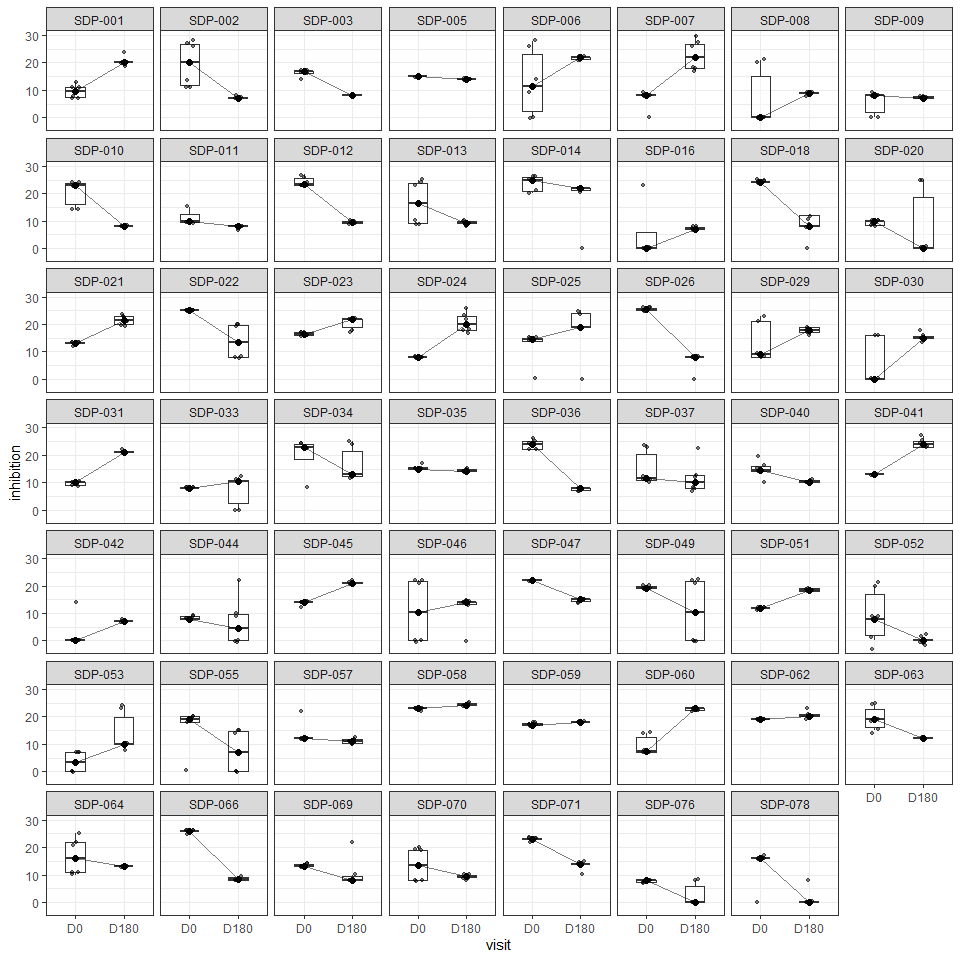

 37: Individual boxplots of inhibition trajectory for E. coli on doxycycline

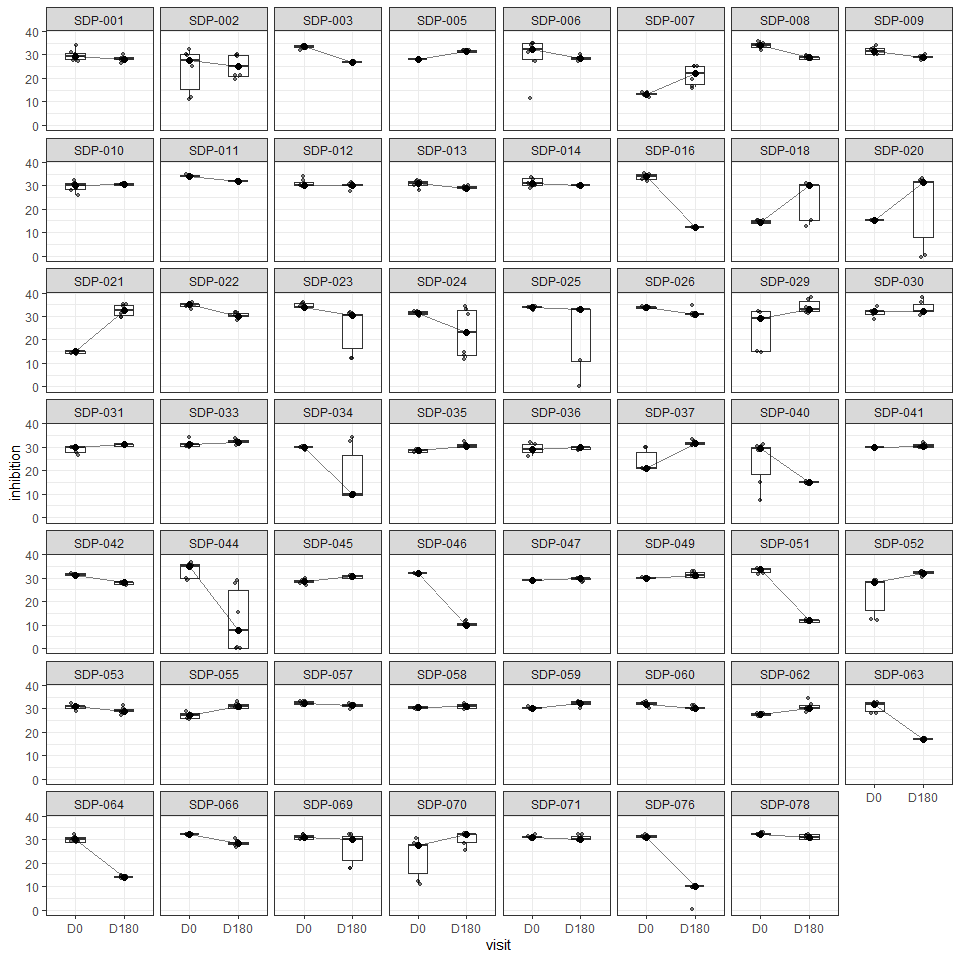

 38: Individual boxplots of inhibition trajectory for E. coli on ceftriaxone

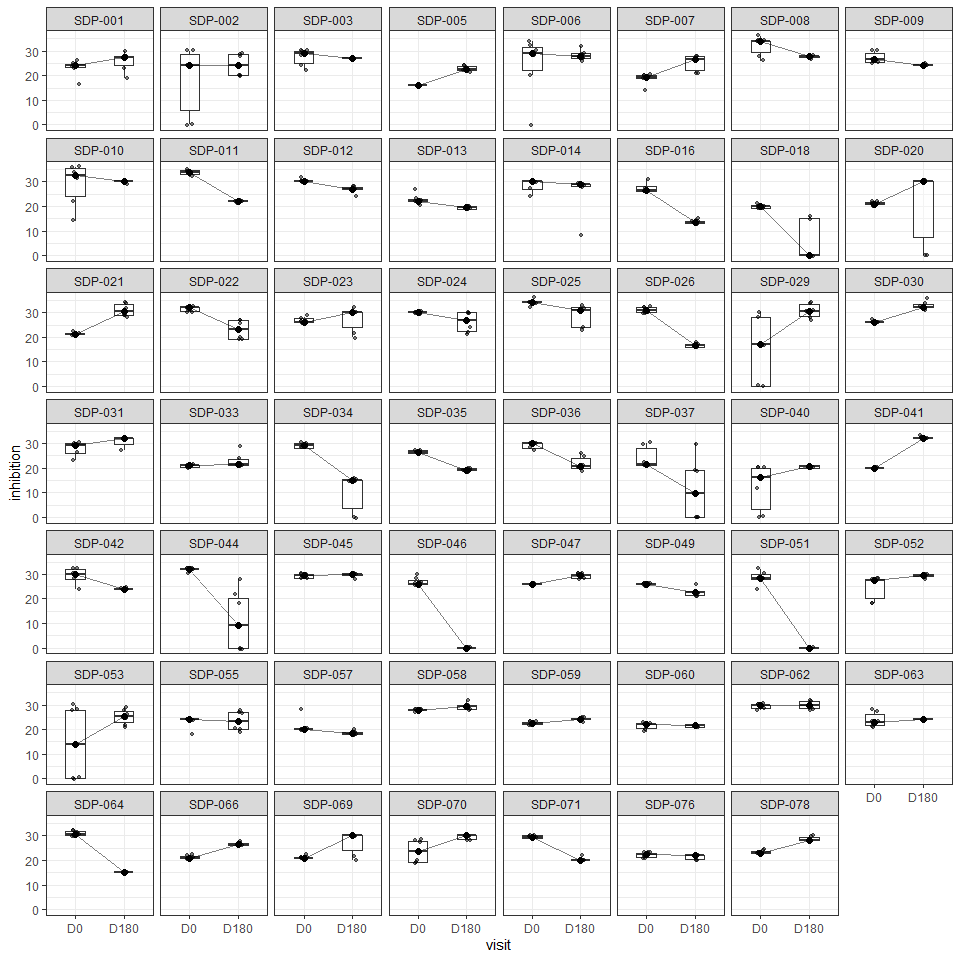

 39: Individual boxplots of inhibition trajectory for E. coli on ciprofloxacin

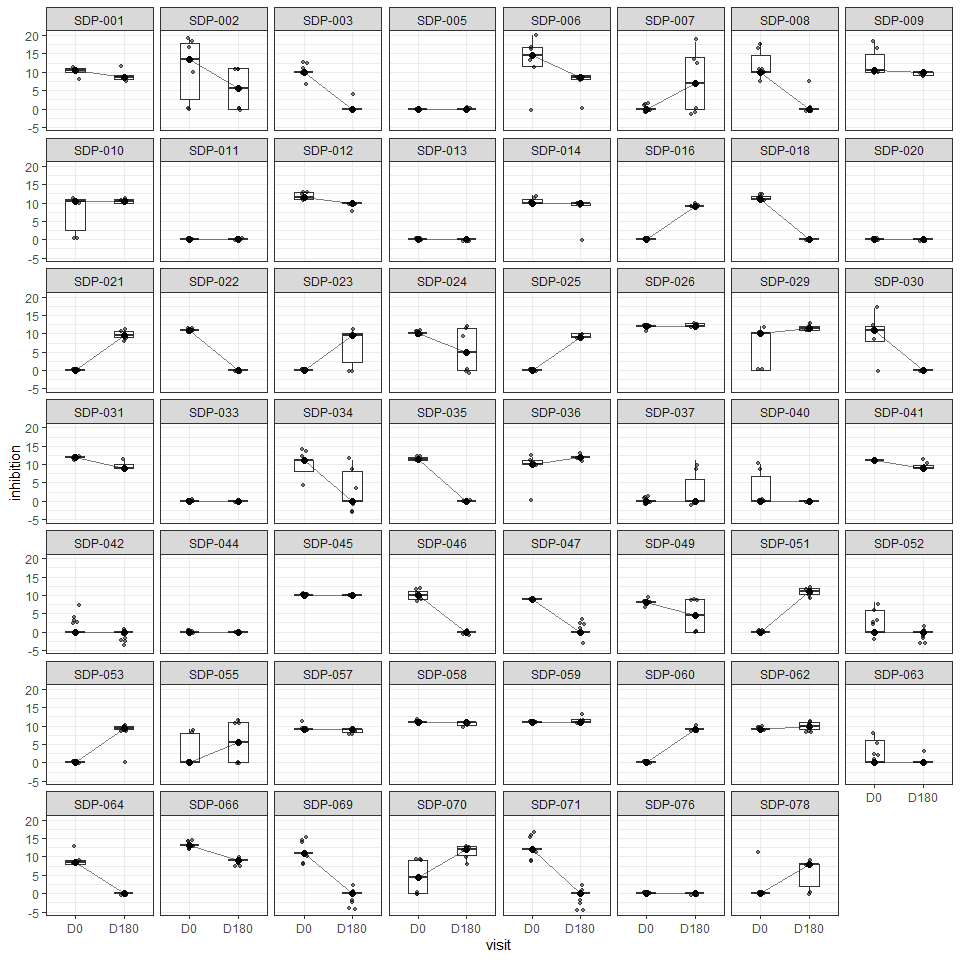

 Individual boxplots of inhibition trajectory for E. coli on azithromycin

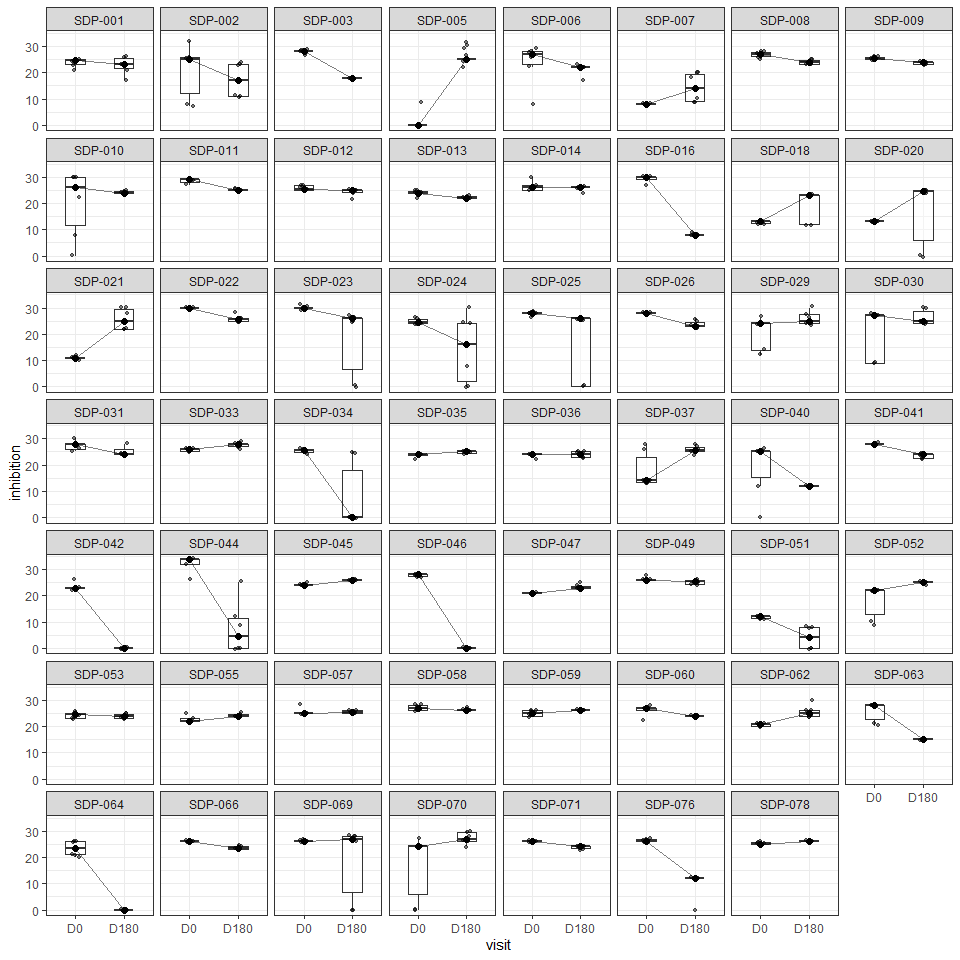

 41: Individual boxplots of inhibition trajectory for E. coli on cefixime

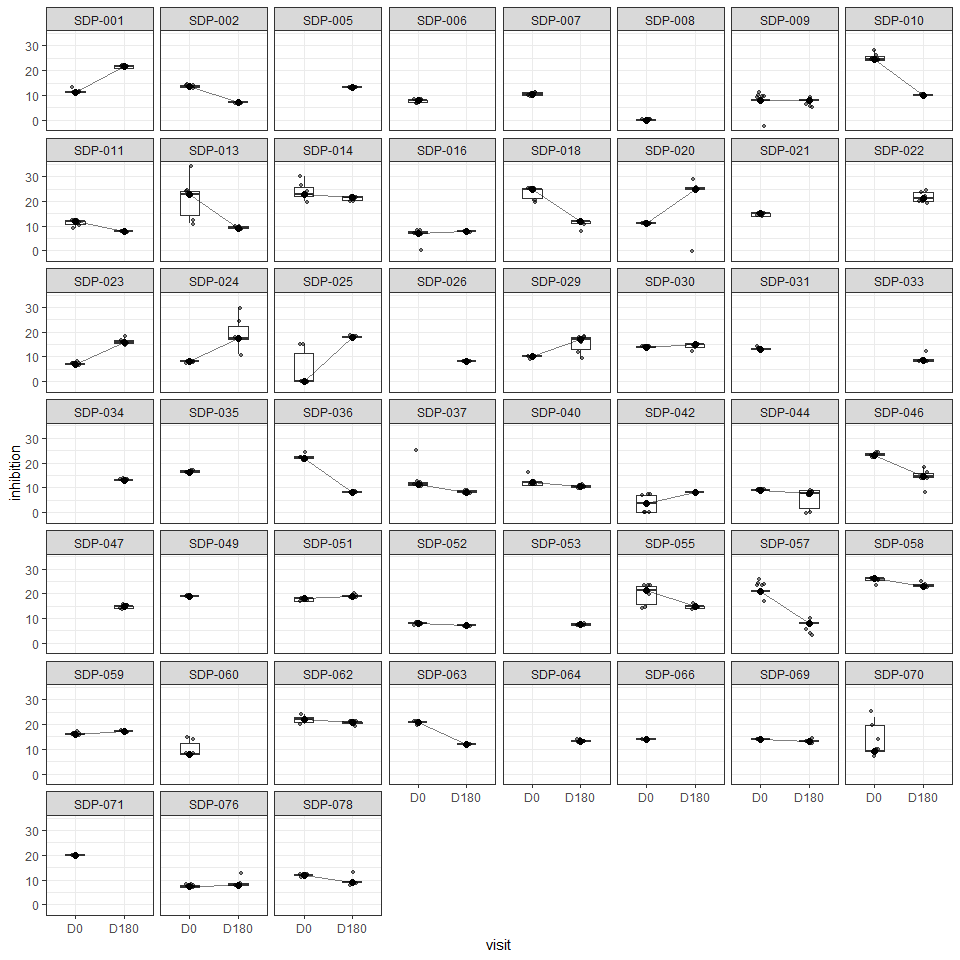

 42: Individual boxplots of inhibition trajectory for Klebsiella pneumoniae on doxycycline

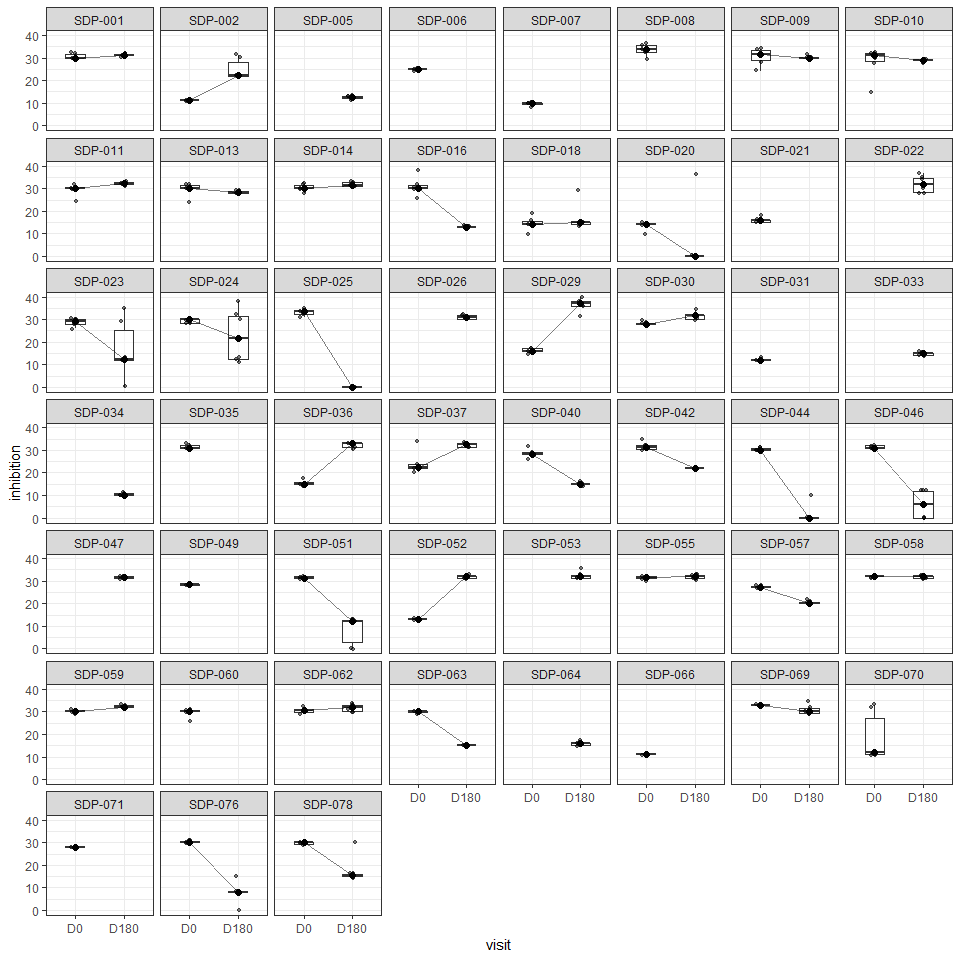

 43: Individual boxplots of inhibition trajectory for Klebsiella pneumoniae on ceftriaxone

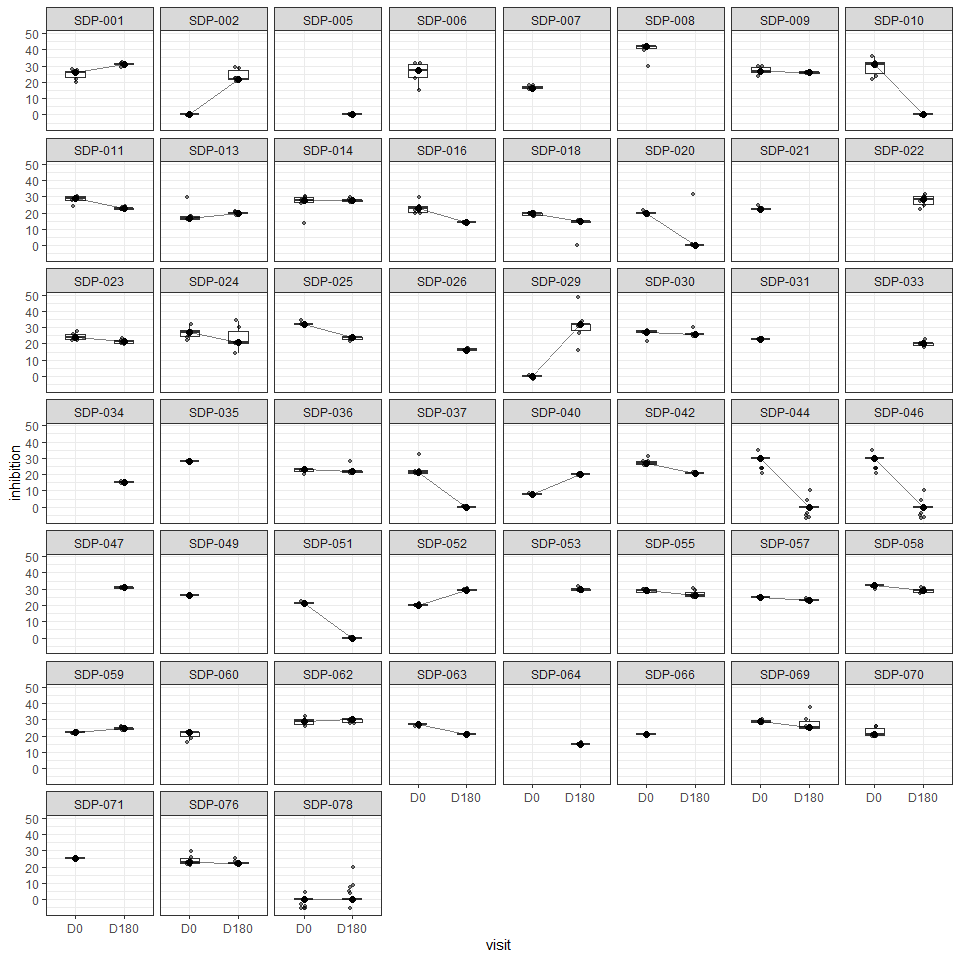

 44: Individual boxplots of inhibition trajectory for Klebsiella pneumoniae on ciprofloxacin

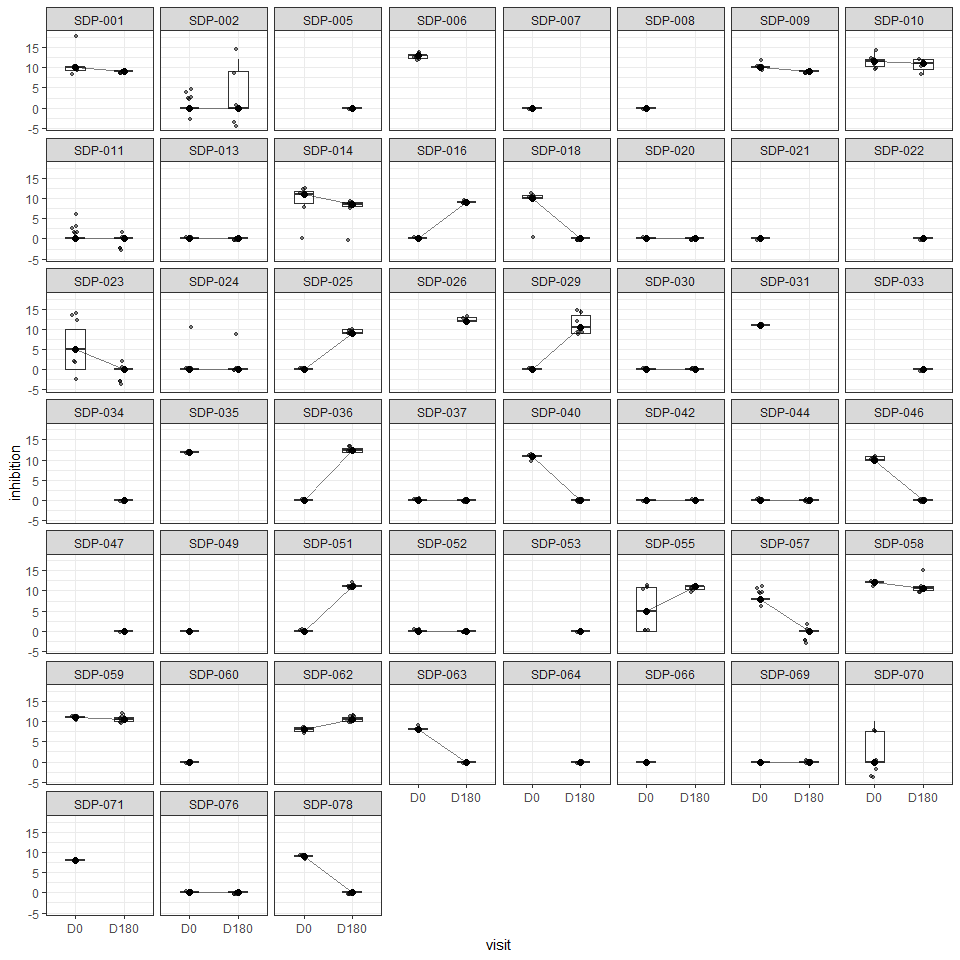

 45: Individual boxplots of inhibition trajectory for Klebsiella pneumoniae on azithromycin

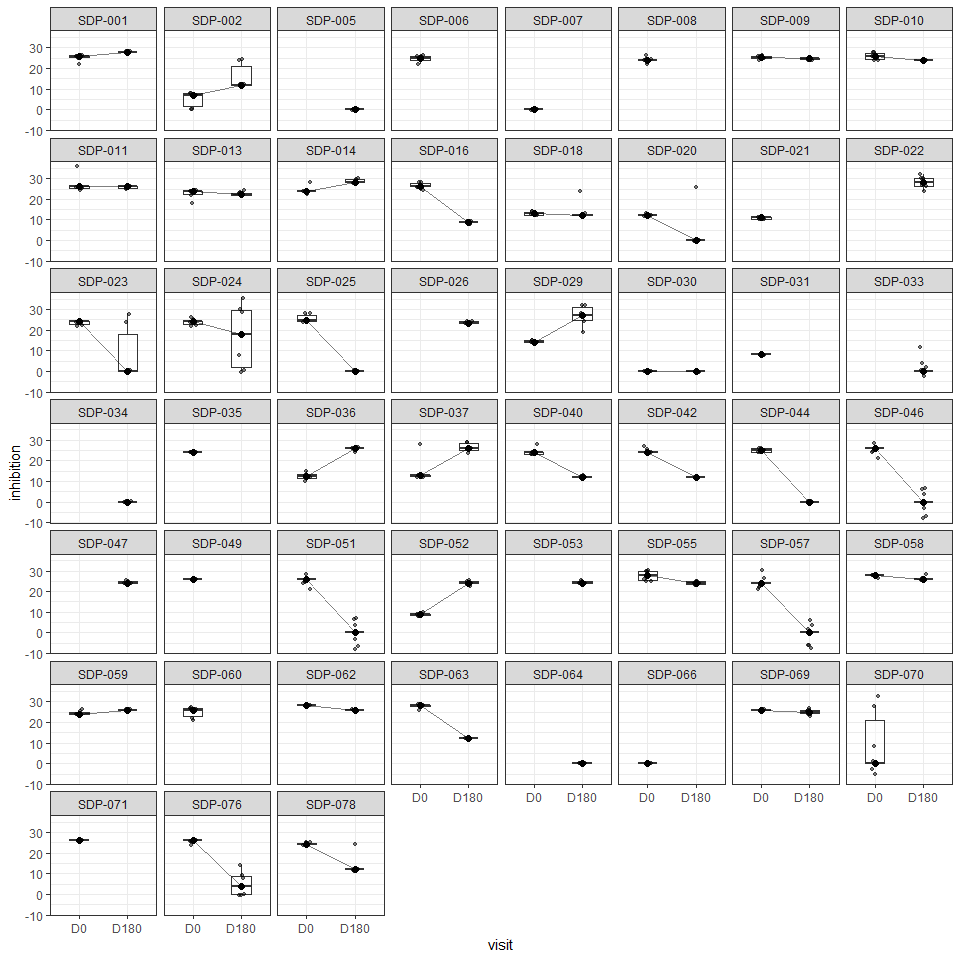

 46: Individual boxplots of inhibition trajectory for Klebsiella pneumoniae on cefixime

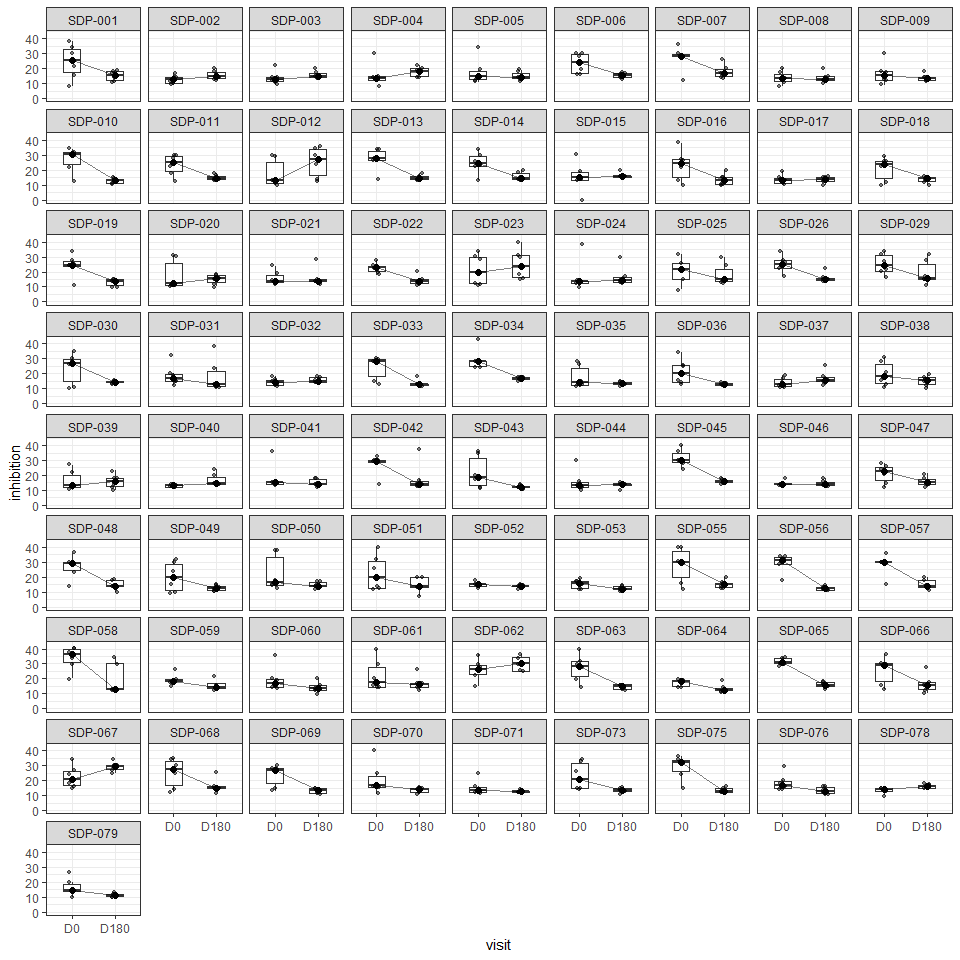

 47: Individual boxplots of inhibition trajectory for oral streptococci on doxycycline

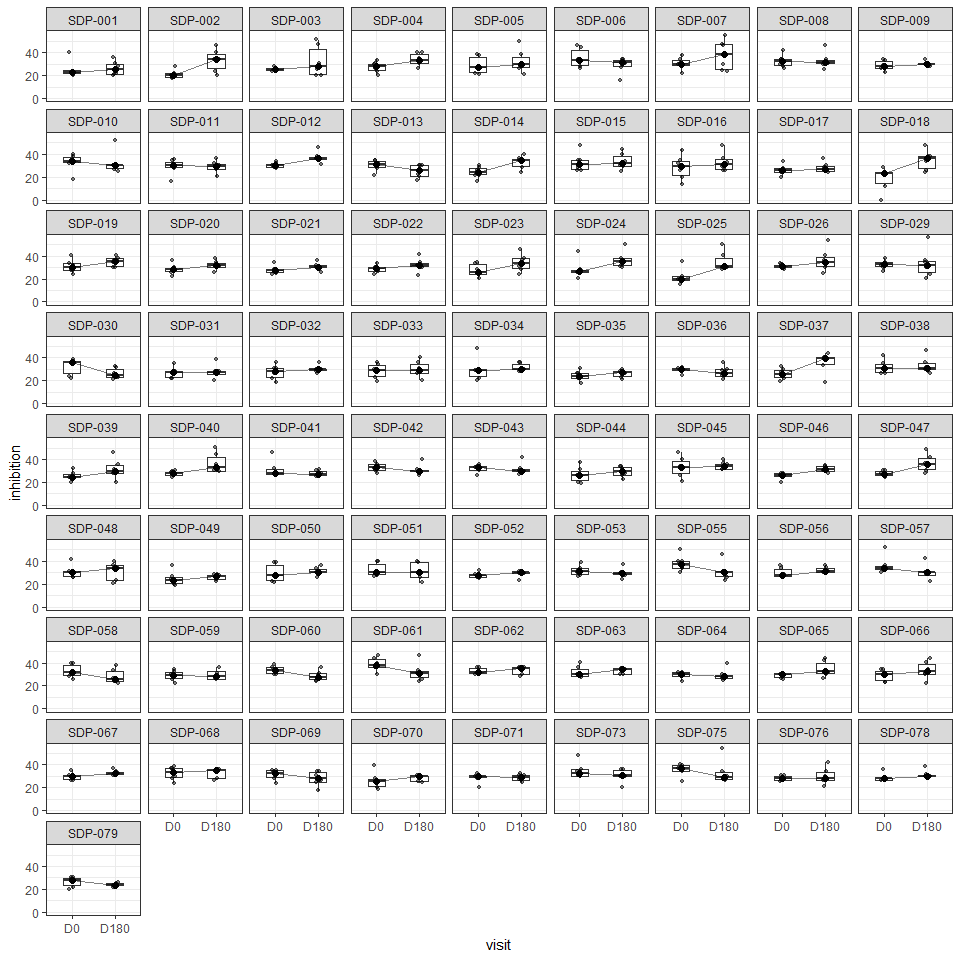

 48: Individual boxplots of inhibition trajectory for oral streptococci on ceftriaxone

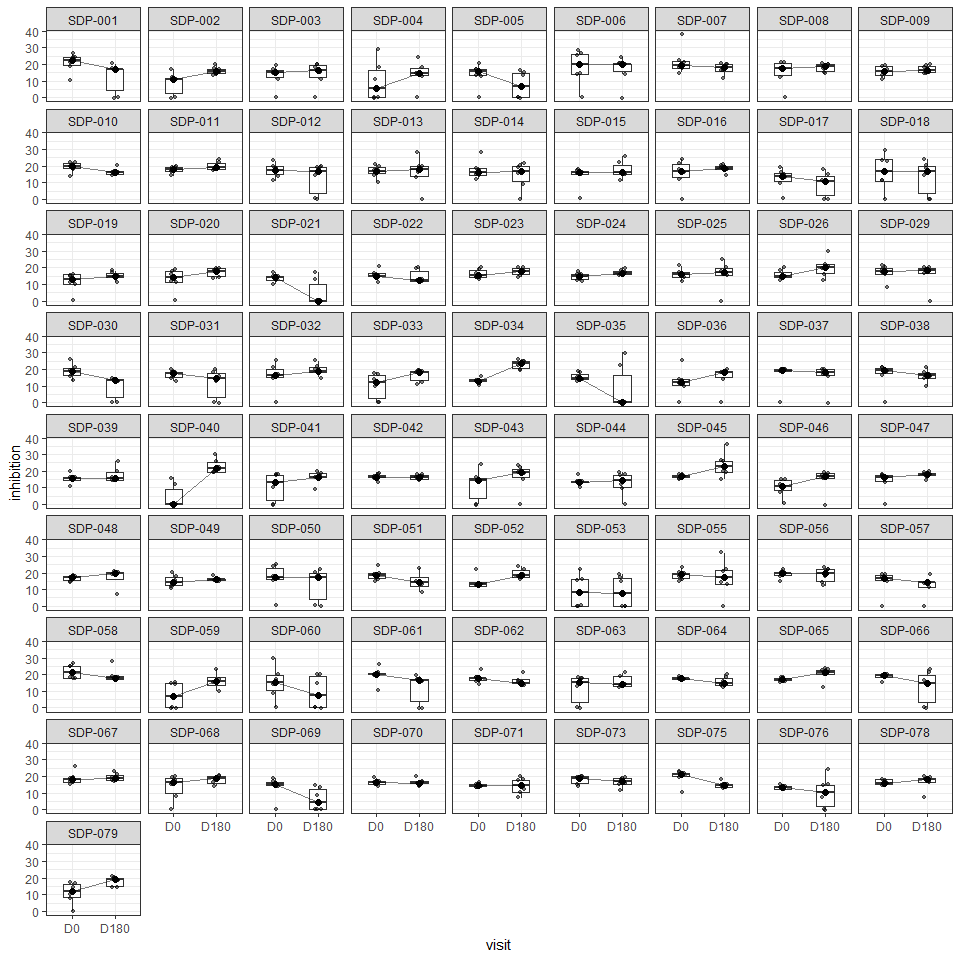

 49: Individual boxplots of inhibition trajectory for oral streptococci on ciprofloxacin

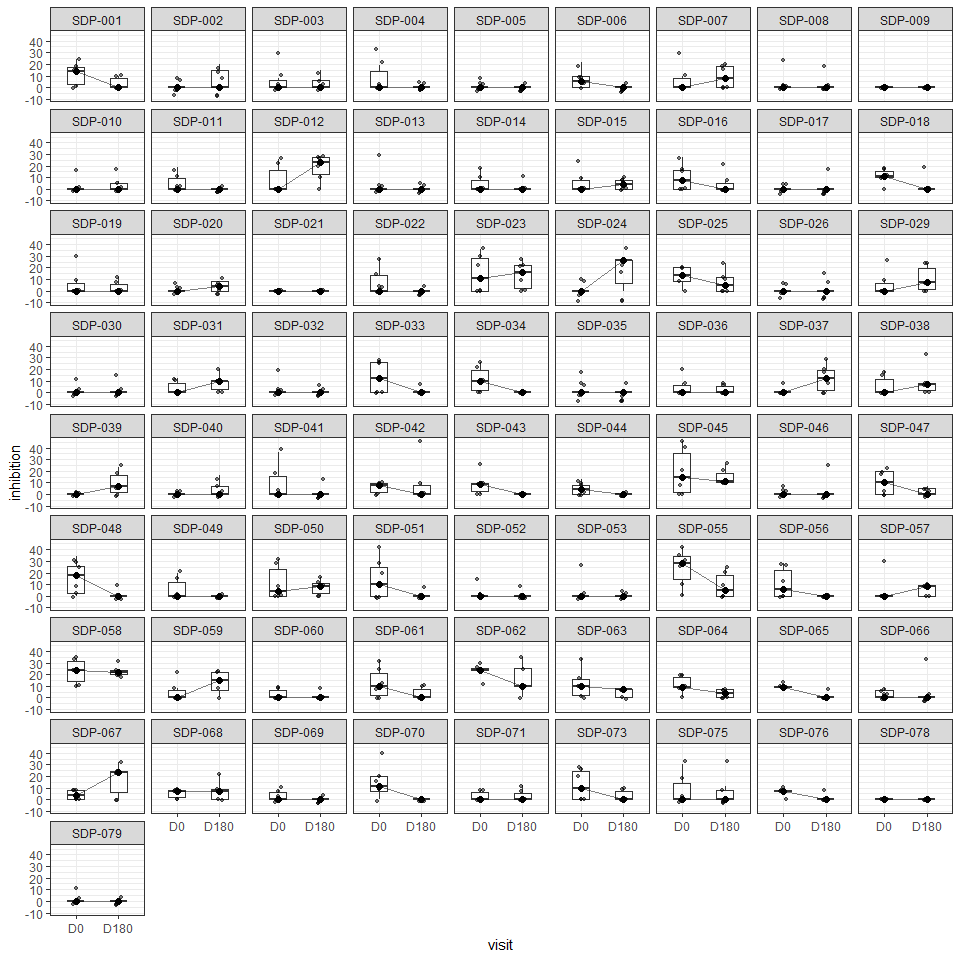

 50: Individual boxplots of inhibition trajectory for oral streptococci on azithromycin

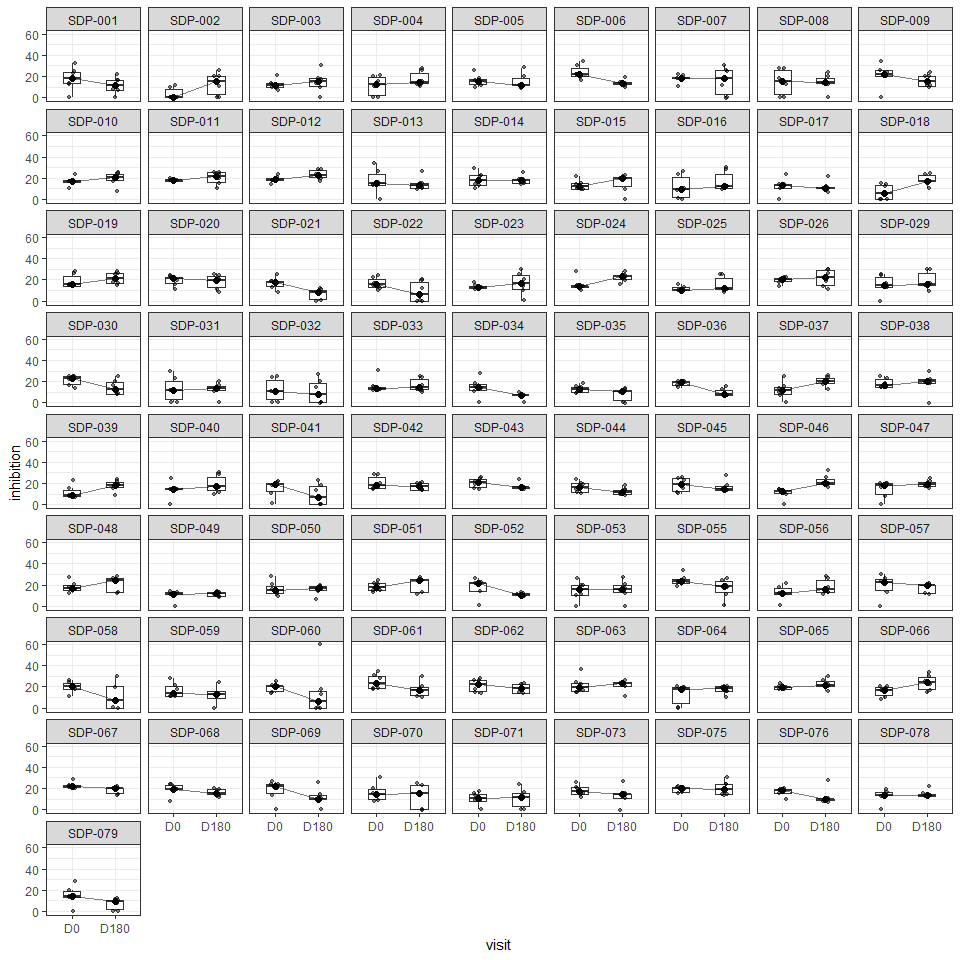

 51: Individual boxplots of inhibition trajectory for oral streptococci on cefixime

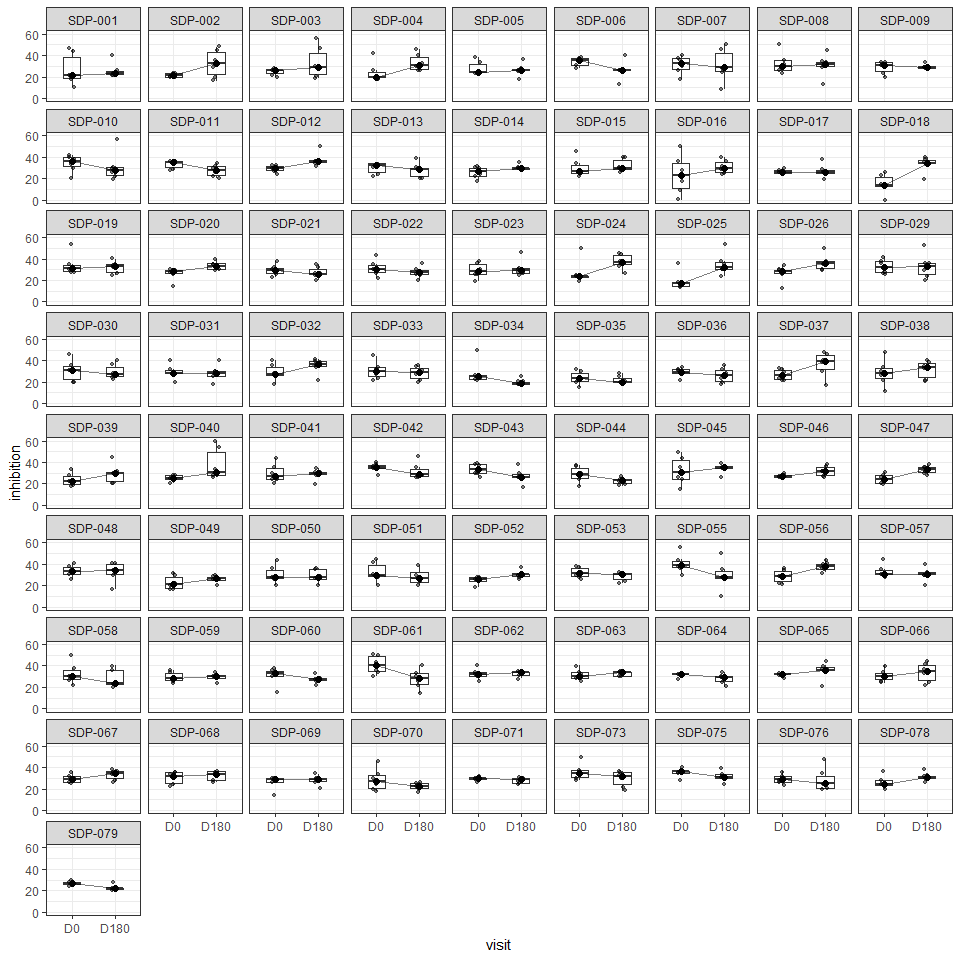

 52: Individual boxplots of inhibition trajectory for oral streptococci on penicilin

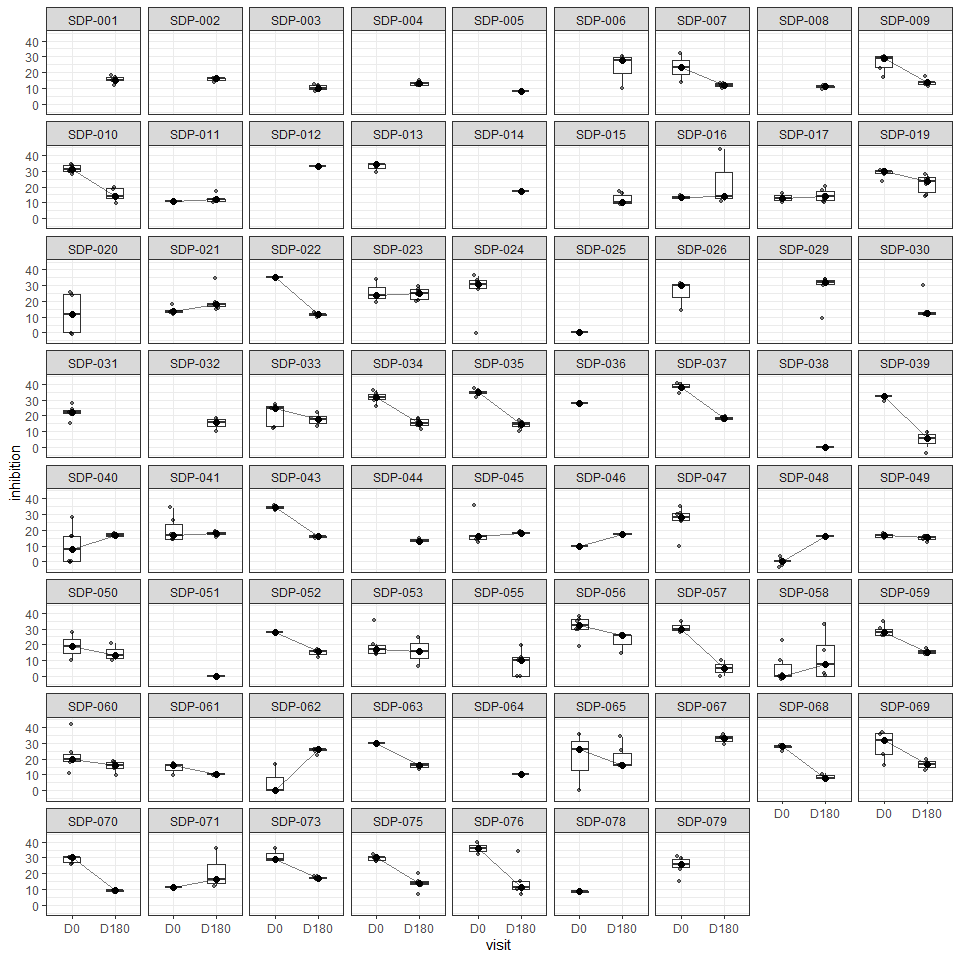

 53: Individual boxplots of inhibition trajectory for Staphylococcus aureus on doxycycline

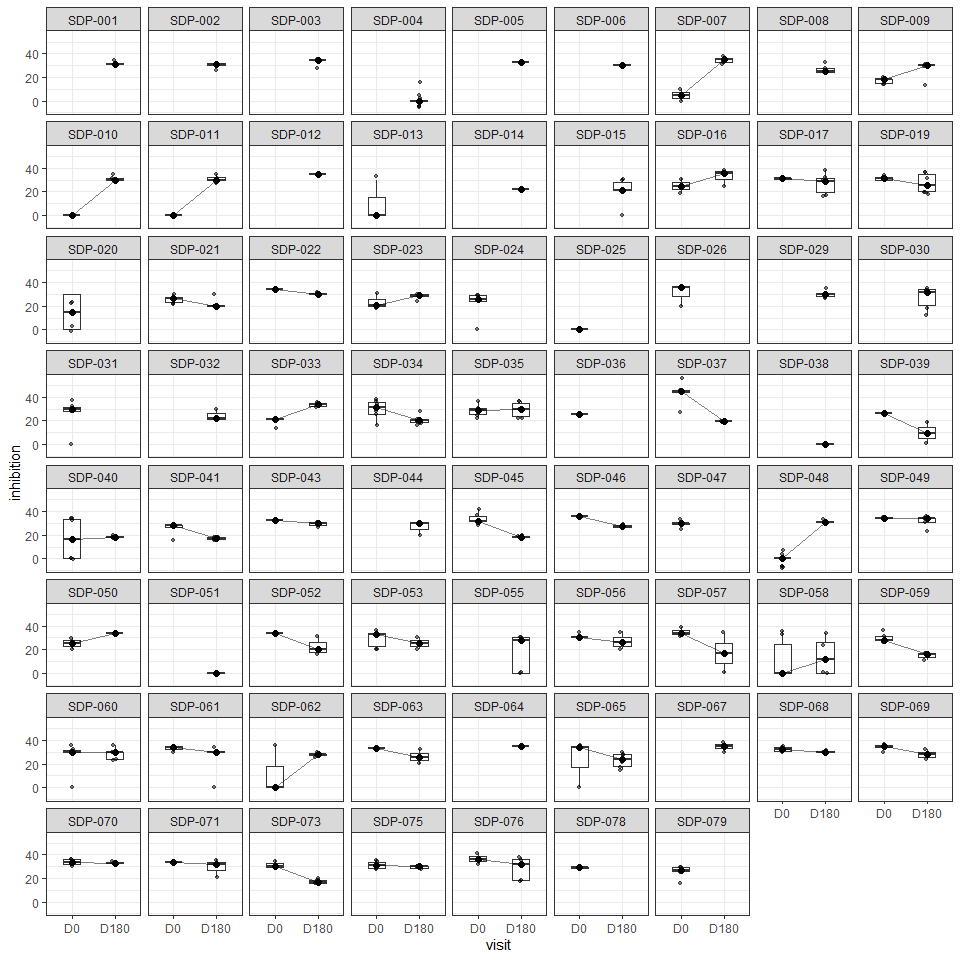

 54: Individual boxplots of inhibition trajectory for Staphylococcus aureus on ceftriaxone

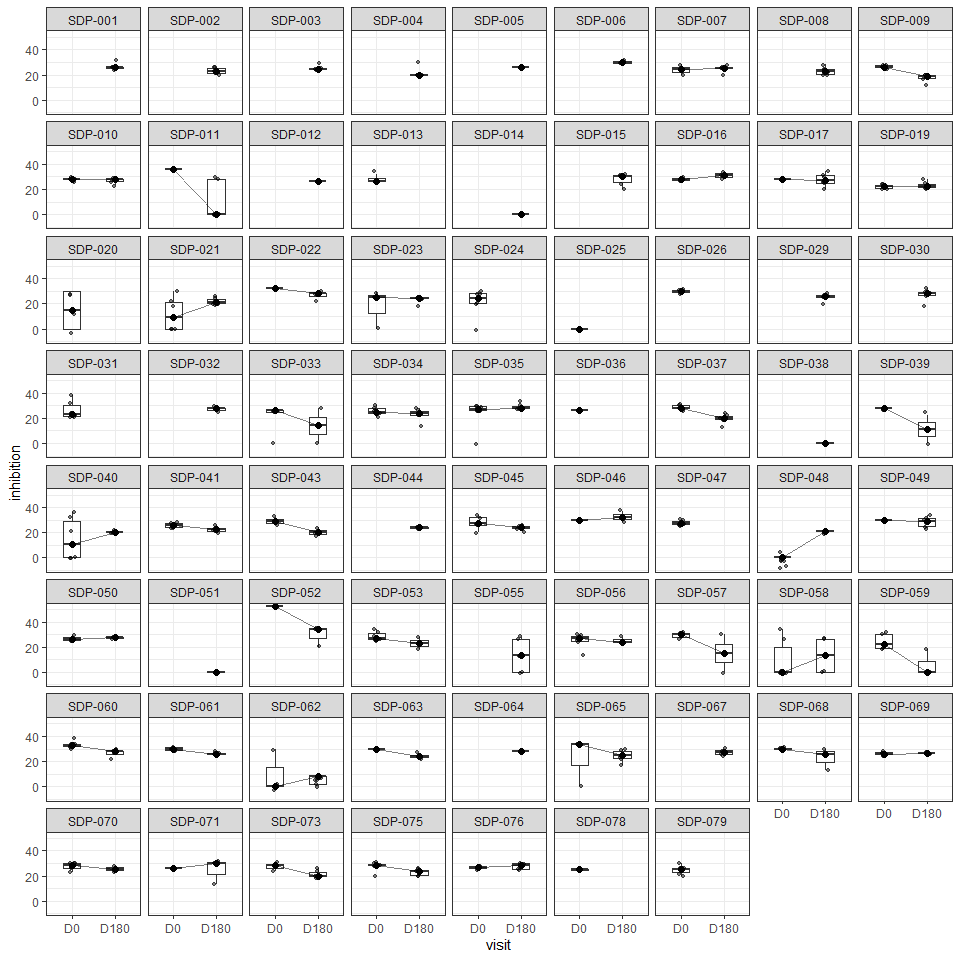

 55: Individual boxplots of inhibition trajectory for Staphylococcus aureus on ciprofloxacin

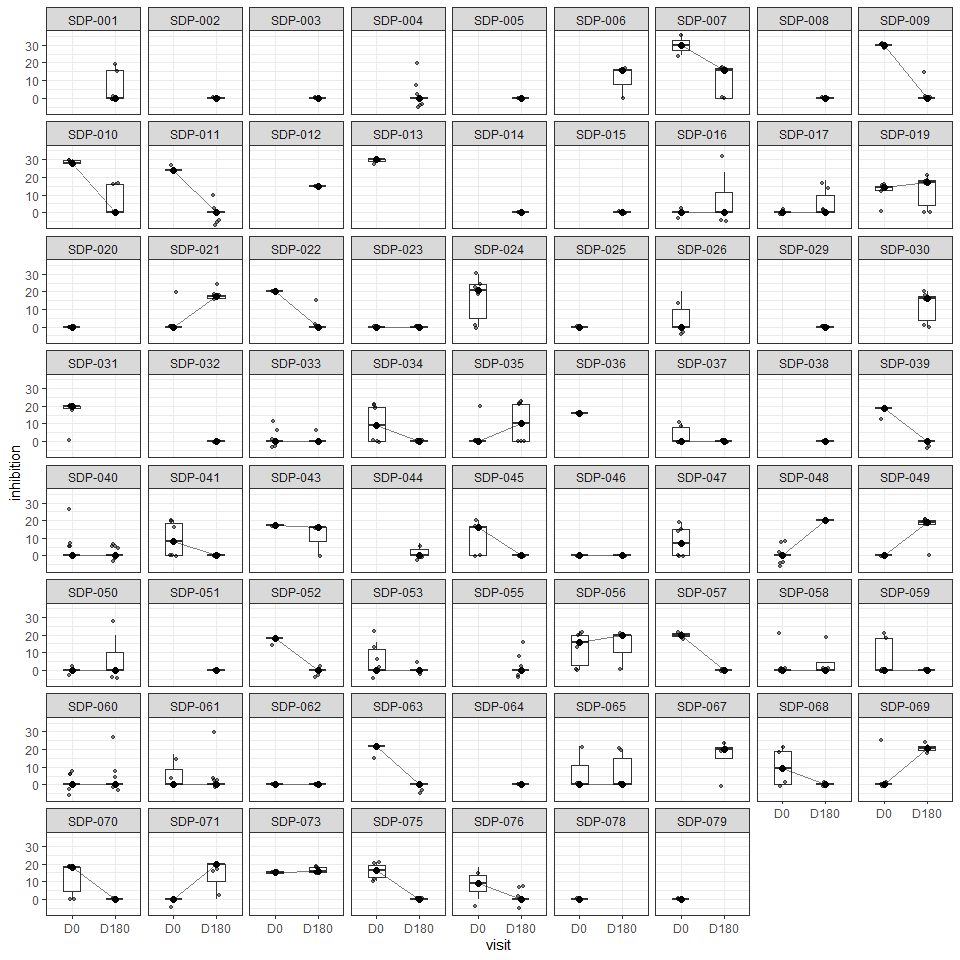

 56: Individual boxplots of inhibition trajectory for Staphylococcus aureus on azithromycin

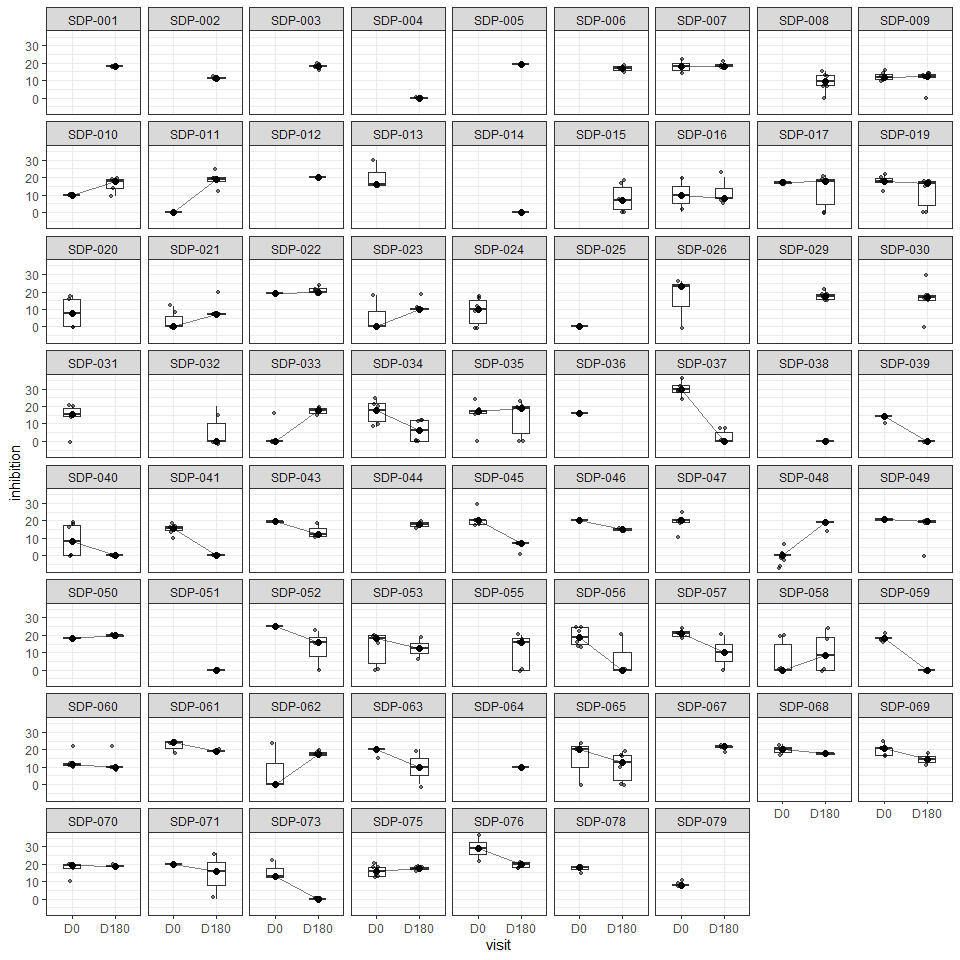

 57: Individual boxplots of inhibition trajectory for Staphylococcus aureus on cefixime

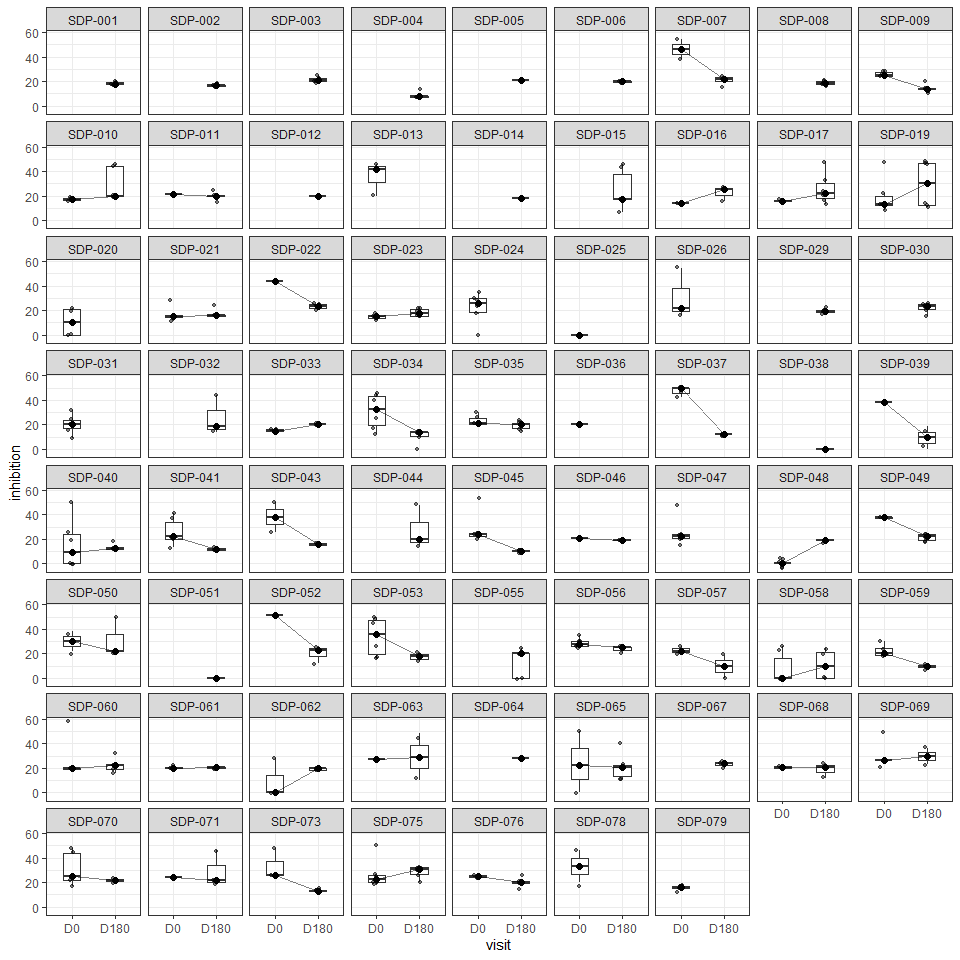

 58: Individual boxplots of inhibition trajectory for Staphylococcus aureus on penicilin

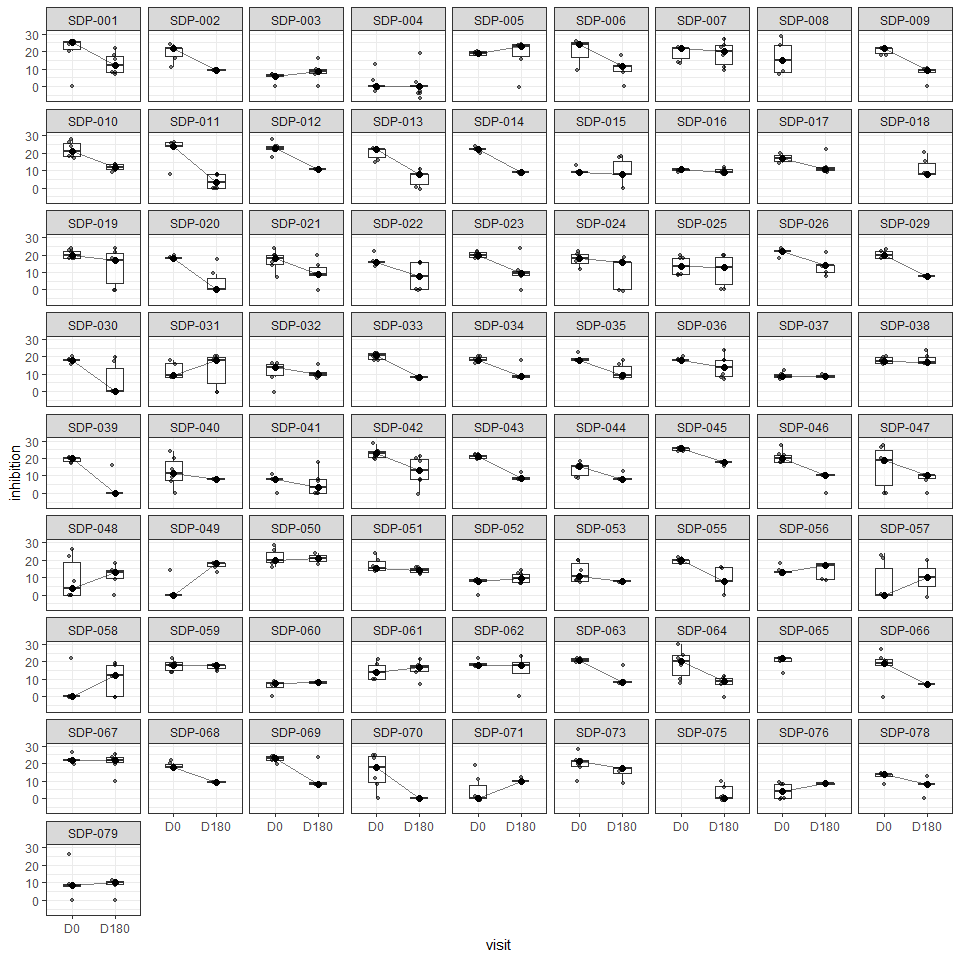

 59: Individual boxplots of inhibition trajectory for Commensal Neisseria on doxycycline

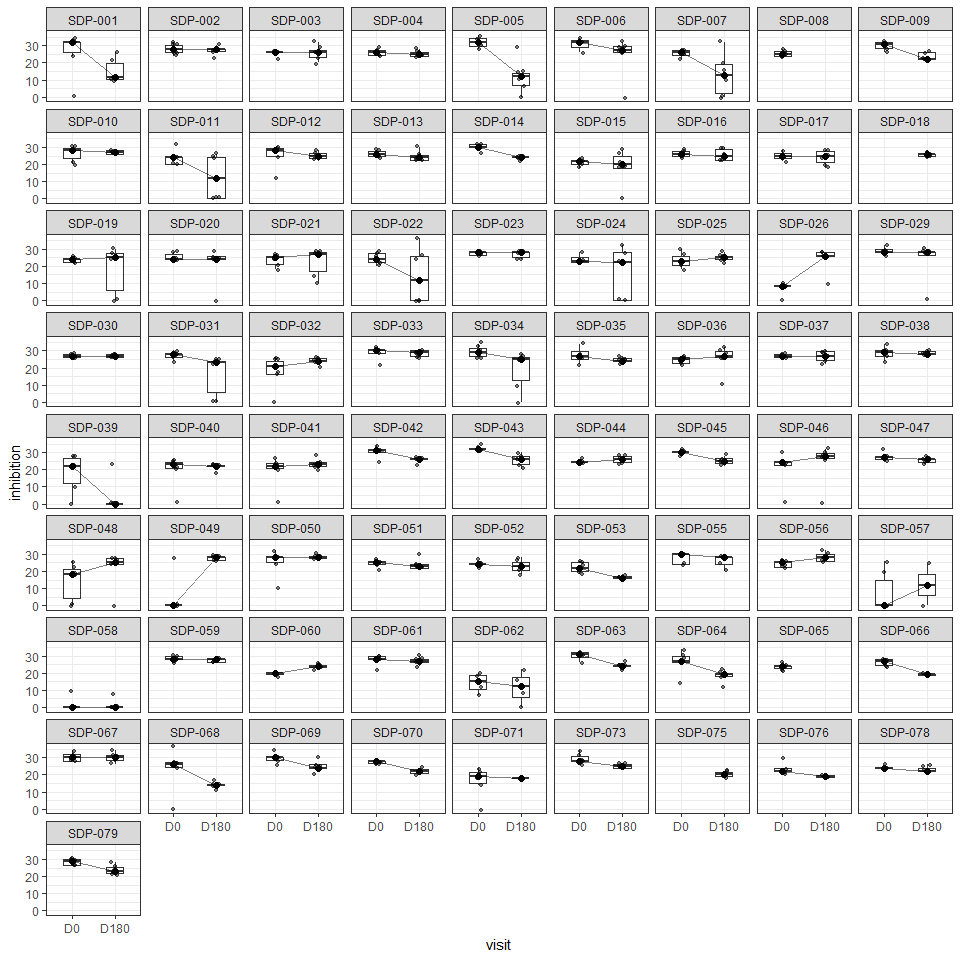

 60: Individual boxplots of inhibition trajectory for Commensal Neisseria on ceftriaxone

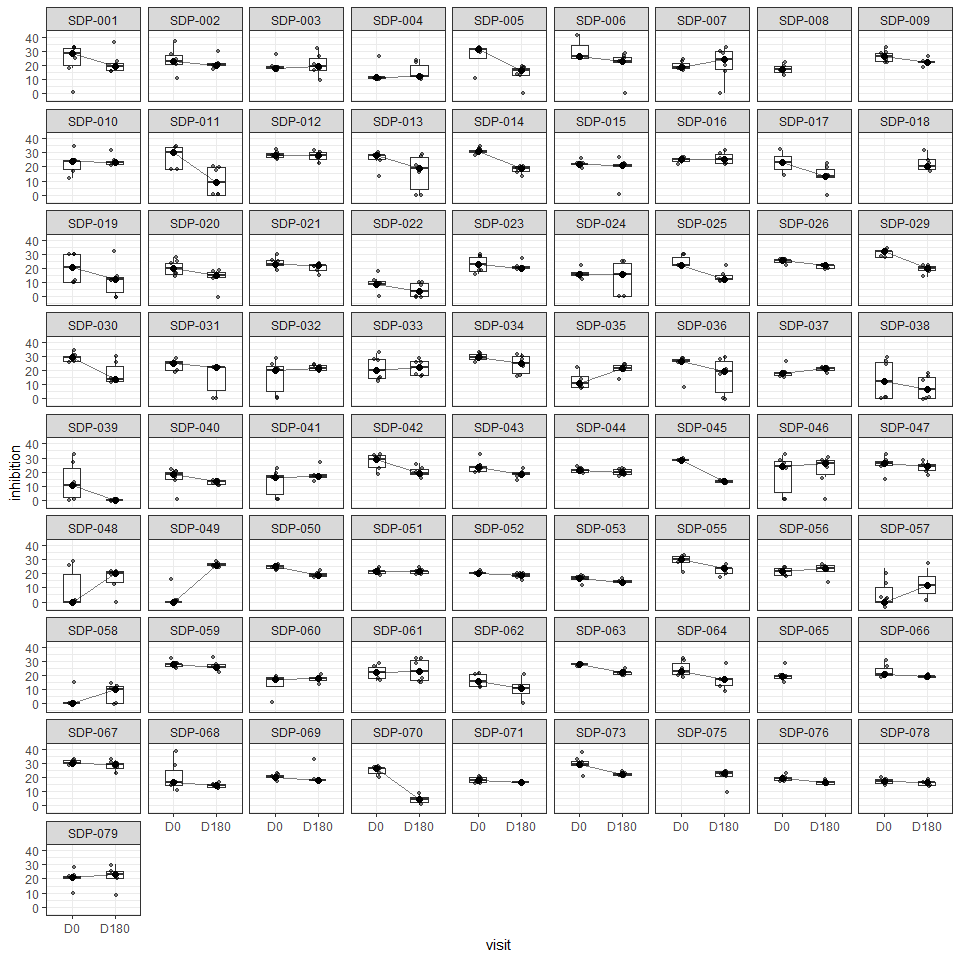

 61: Individual boxplots of inhibition trajectory for Commensal Neisseria on ciprofloxacin

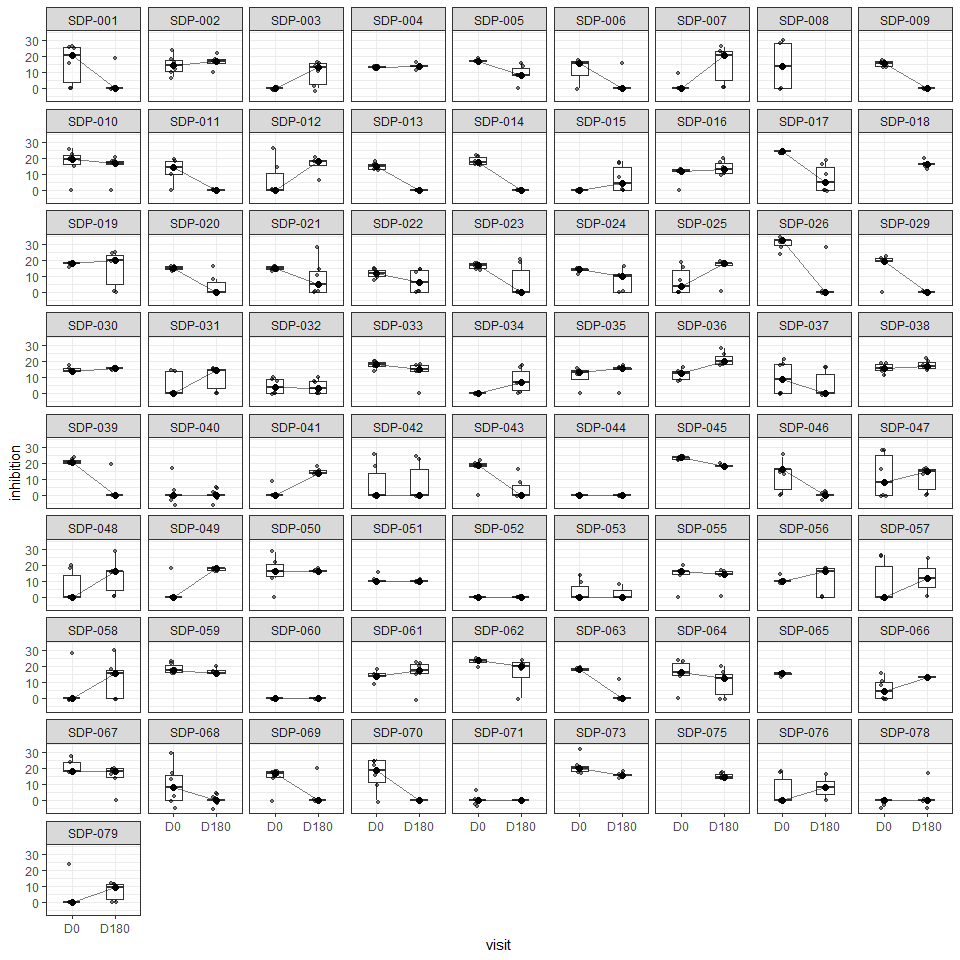

 62: Individual boxplots of inhibition trajectory for Commensal Neisseria on azithromycin

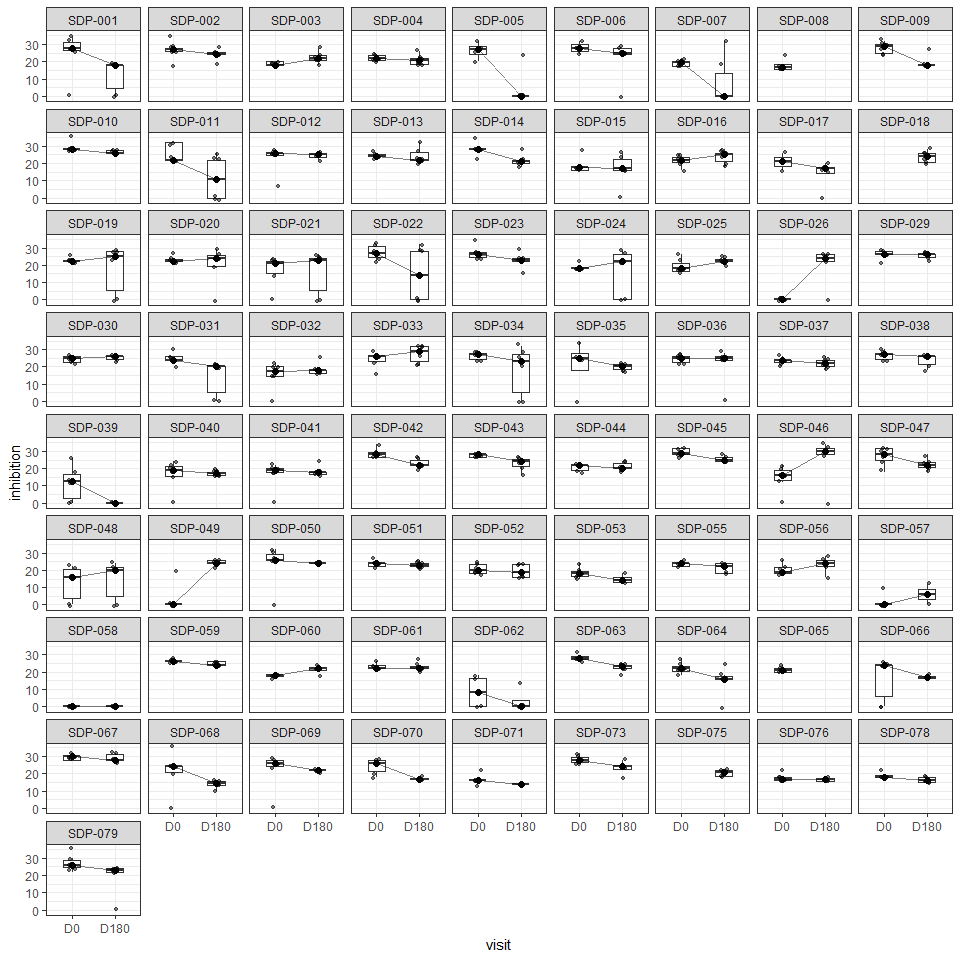

 63: Individual boxplots of inhibition trajectory for Commensal Neisseria on cefixime

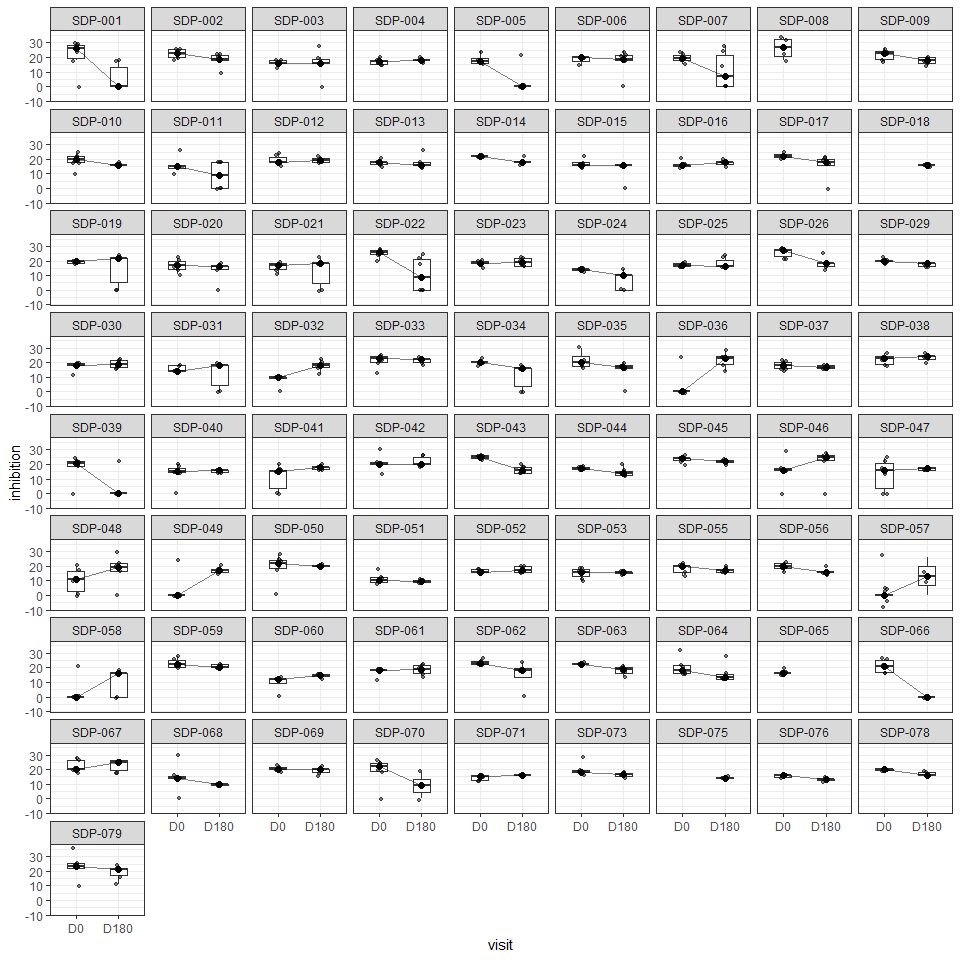

 64: Individual boxplots of inhibition trajectory for Commensal Neisseria on penicilin
